# Association between anticholinergic burden and dementia in UK Biobank

**DOI:** 10.1101/2021.08.04.21261330

**Authors:** J. Mur, T.C. Russ, S. R. Cox, R. E. Marioni, G. Muniz-Terrera

## Abstract

Previous studies on the association between the long-term use of anticholinergic drugs and dementia report heterogenous results. This variability could be due to, among other factors, different anticholinergic scales used, and differential effects of distinct classes of anticholinergic drugs. Here, we use 171,775 participants of UK Biobank with linked GP prescription records to calculate the cumulative annual anticholinergic burden (ACB) and ascertain dementia diagnoses through GP- and inpatient records. We then compare 13 anticholinergic scales and anticholinergic burden (ACB) due to different classes of drugs in their association with dementia. We find dementia to be more strongly predicted by ACB than by polypharmacy across most anticholinergic scales (standardised ORs range: 1.027-1.125). Furthermore, not only the baseline ACB, but the slope of the longitudinal trajectory of ACB (HR=1.094; 95% CI: 1.068-1.119) is predictive of dementia. However, the association between ACB and dementia holds only for some classes of drugs – especially antidepressants, antiepileptics, and high-ceiling antidiuretics. Moreover, we do not find a clear relationship between reported anticholinergic potency and dementia risk. The heterogeneity in findings on the association between ACB and dementia may in part be due to different effects for different classes of drugs. Future studies should establish such differences in more detail and further examine the practicality of using a general measure of anticholinergic potency as it relates to the risk of dementia.

## Introduction

The number of people with dementia is predicted to increase in the UK by 50% from the year 2016 to 2040 and worldwide from 50 million today to 152 million in 30 years^1^. Considering the lack of treatment options, the specification of risk factors to reduce the incidence of the disease is crucial. It is estimated that ∼40% of risk factors for dementia are preventable^1^ and that the decreases in the incidence of dementia in some countries are partly attributable to reductions in some of these risk factors^2^.

Anticholinergic drugs block muscarinic acetylcholine receptors in the nervous system, which are important in the innervation of brain areas involved in cognitive function, and in the pathophysiology of Alzheimer’s disease^3^. Due to their mechanism of action, sustained use of these medicines might impair cognitive function later in life. Anticholinergic burden (ACB) – a measure of anticholinergic drug use – has indeed been linked to an increased risk of delirium, cognitive impairment, and dementia in older people^4^. Recent studies have focused on the long-term effects of anticholinergic drugs when taken before advanced age. For certain anticholinergic drugs, these studies report an increased rate of dementia following their use decades before the diagnosis^5,6^. This suggests a potential for ACB as a marker for cognitive decline, or as a causative risk factor. In other words, ACB could be indicative of comorbidities that themselves affect cognition or could, through the drugs’ mechanism of action, contribute to cognitive decline as an independent risk factor. However, the status of anticholinergic medication in dementia prevention is unclear, as several recent reviews on the topic report heterogenous findings^7–9^.

The variability in previous findings can be partly explained by differences in study design, the characteristics of the samples, the covariates in the models, and the choice of anticholinergic scales that assign drugs their anticholinergic potency. There is no widely accepted procedure to score anticholinergic potency1^0^, and anticholinergic scales were constructed in distinct regions, contexts, and validated in different samples. This leads to poor agreement among anticholinergic scales and uncertainty when choosing among them^11,12^.

Additionally, the association between ACB and dementia may be small. Considering that the samples in previous studies often do not include more than a few thousand participants^8^, they possibly lack the power to detect these differences. Moreover, the proposed harmful effect of anticholinergic drugs on cognition perhaps arises only in some classes of drugs. Recent studies exploring class-based associations report effects especially for antidepressants, urological drugs, and antipsychotics^5,6^. Thus, broad recommendations of (de-)prescribing for any anticholinergic drug for any patient might not be appropriate. This is especially the case since drugs are prescribed to manage underlying conditions that themselves decrease the quality of life and in cases when drug alternatives that exhibit fewer side effects are unavailable.

To elucidate the proposed association between ACB and dementia, well-powered replication studies and detailed inspections of the effects of different anticholinergic scales and drug classes are necessary. Here, using data from a large population-based biobank (UK Biobank, n=230,000), we compare different anticholinergic scales in their propensity to predict dementia and explore the effect of within-person trajectories in ACB on the risk of dementia. We then attempt to replicate the results from other studies^5,6^ that linked the use of some classes of anticholinergic drugs to an increased risk of dementia and that investigated the risk for dementia at different latencies from the period of anticholinergic use.

## Methods

### Hypotheses

We predicted ACB to be positively associated with dementia across anticholinergic scales and the association to be stronger than the association between dementia and polypharmacy. Next, we expected the precision of the association between ACB and dementia to increase within scales in the following sequence: count-based version, value-based version, dosage-based version. We also anticipated the evolution of ACB over time to be positively associated with dementia. Finally, based on previous studies^5,6^, we hypothesised that ACB due to antidepressants, antihistamines, antiepileptics, urological-, and antipsychotic drugs will show a positive association with dementia. The association between other classes of drugs and dementia, and the analysis of latencies between ACB and dementia was not based on prior hypotheses.

### Sample

UK Biobank is a prospective study of >500,000 participants that were recruited across the UK from 2006-2010^13^. For ∼230,000 of these participants, primary-care electronic prescription entries are available until September 2017. The entries contain the drugs prescribed, dates of prescriptions, and Read-codes (https://isd.digital.nhs.uk) that act as dictionaries for medicines. Diagnoses were obtained by combining primary care- and inpatient hospital records for each participant and – in cases of multiple entries for a disorder – retaining the earliest record (**Suppl. Table 1**).

### Anticholinergic burden and drug class

Eleven anticholinergic scales^14,15,24,16–23^ were chosen as previously identified (Mur et al., DOI:10.1101/2020.10.16.20213884) and two^25,26^ were identified through a recent systematic review^27^ (**Suppl. Table 2**). One scale^22^ was modified to include newer drugs as before^28^; for two scales^15,17^, updated versions were used (Aging Brain Care, 2012; Carnahan, 2014, personal communication on 21.10.2019). For one scale^19^, drugs classified by the authors as having “improbable anticholinergic action”, were assigned an anticholinergic burden of 0.5 (between “no anticholinergic potency” and “weak anticholinergic potency”).

Using the British National Formulary (https://bnf.nice.org.uk), brand names of anticholinergic drugs in the sample were substituted with generic names. Combination prescriptions containing several anticholinergic compounds were separated into multiple entries, each containing a single anticholinergic compound.

Each prescription was assigned anticholinergic scores based on the ratings from anticholinergic scales. Prescriptions of drugs with ophthalmic, otic, nasal, or topical routes of administration were assigned an anticholinergic score of 0, as before^21–24^. For each scale, ACB was estimated by three separate means. First, the total yearly number of anticholinergic drugs was determined (count-based scale). Second, each drug was assigned the anticholinergic value as listed in the anticholinergic scale and the values were summed for each year (value-based scale). Third, a standardised dosage was calculated for each prescription by dividing the prescribed dose by the defined daily dose (DDD, https://www.whocc.no) and then it was multiplied by the anticholinergic score (dosage-based scale). For the dosage-based scale, years in which any anticholinergic prescription was missing information on dosage, were removed for that participant (751 observations, 0.43% of the sample). The quantity of drugs (e.g., number of tablets in a prescription) was not accounted for in this calculation due to the high number of missing values (∼25% of prescriptions). Each drug was assigned to a class based on the Anatomical Therapeutic Chemical (ATC) Classification System (https://www.whocc.no) (**Suppl. Table 3**), and to a group of anticholinergic potency (groups 0, 0.5, 1, 2; a higher value indicates a greater presumed anticholinergic potency) according to the anticholinergic scale^19^ with the strongest effect size for predicting dementia (see below).

### Covariates and statistical analysis

The predictor in most models was the cumulative ACB in year 0 (the sum of anticholinergic scores of prescriptions for a participant). Due to the low ascertainment of prescriptions in the early years of sampling (Mur et al., DOI:10.1101/2020.10.16.20213884), year 0 was for each participant defined as the first full year of having been included in the prescriptions’ register after the year 1999.

Because the rate of dementia increases with age, participants younger than 60 years at time of diagnosis or at the end of the prescriptions sampling period (30.06.2020) – whichever came first – were excluded from the analyses. Additionally, participants, who before year 0 or within a year after year 0, had been diagnosed with dementia or prescribed with a cholinesterase inhibitor (donepezil, galantamine, or rivastigmine) or memantine were excluded from the analyses. We also excluded participants diagnosed at any point with Parkinson’s disease, Huntington disease, Creutzfeldt-Jacob disease, or multiple sclerosis. Finally, the prescribing period after the year 2015 was incomplete (Mur et al., DOI:10.1101/2020.10.16.20213884) and was removed. The data cleaning process is described in **Suppl. Figure 1**.

Models were adjusted for sex (reference: female), data provider (region-specific providers of prescriptions: The Phoenix Partnership (TPP) England, Vision England (reference), Vision/EMIS Health Scotland, Vision/EMIS Health Wales), education (binary; reference: no graduate degree), socioeconomic deprivation based on census data (scale range:-12-12; range in sample:-6.3-7.4; bigger number indicates greater deprivation)^29^, body mass index (BMI in kg/m^2^, categorised: <18.5, 18.5-25 (reference), 25-30, 30-35, 35-40, >40), self-reported smoking status (smoker, non-smoker (reference), former smoker), self-reported alcohol consumption frequency (daily or almost daily (reference), three or four times a week, once or twice a week, one to three times a month, only on special occasions, never), self-reported physical activity (mild (reference), moderate, strenuous)^30^, number of comorbidities (number of all unique diagnosis codes) by year 0, depression by year 0 (reference: no depression), stroke by year 0 (reference: no stroke), diabetes by year 0 (reference: no diabetes), hypercholesterolemia by year 0 (reference: no hypercholesterolemia), hypertension by year 0 (reference: no hypertension), *APOE*-carrier status (reference: ε2), and polypharmacy. The latter was determined separately for each anticholinergic scale by subtracting the yearly number of anticholinergic drugs according to that scale from the total yearly drug count. Diagnoses were ascertained using both hospital-(UK Biobank data fields 41270 and 41270) and primary-care records (UK Biobank data field 42040). *APOE* genotype was determined based on the nucleotides at SNP positions rs239358 and rs7412; *APOE*-carrier status was denoted as ε3 for participants with the ε3/ε3 haplotype, ε2 for participants with the ε2/ε2 or ε2/ε3 haplotype, and ε4 for participants with the ε3/ε4 or ε4/ε4 haplotype.

For the association between ACB and dementia, Cox proportional hazards models were used, and effects are expressed as hazard ratios (HRs) with accompanying 95% confidence intervals (CIs). For studying time-to-event latencies, logistic regression was used, and effects are expressed in odds ratios (ORs). The association between the longitudinal evolution of ACB and dementia accounted for the competing risk of death and was assessed with the joint model for longitudinal and time-to-event data using the R library *JM*^31^. To compare anticholinergic scales, separate models were run for each scale. Additionally, two models were run with polypharmacy as the predictor. For all other analyses using only a single anticholinergic scale, the value-based scale by Durán et al. (2013)^19^ was used, as it exhibited the strongest association with dementia. Models for which ACB was the main predictor were additionally controlled for polypharmacy. The model for which polypharmacy was the main predictor, was run in two steps. First, we controlled for the covariates described above except for polypharmacy. Second, we additionally controlled for the total number of anticholinergic drugs (according to any anticholinergic scale).

Numerical values three or more standard deviations beyond the mean were defined as outliers and removed from the analytical sample prior to analysis. Due to zero-inflation for ACB, the number of prescriptions, and the number of comorbidities, null-values were removed before calculating means and standard deviations for outlier removal for these variables. Cases with missing values were removed prior to analysis and constituted up to 16.9% of the sample, depending on the model. When exploring the ACB attributable to different drug classes, only drug classes were included for which at least ten prescriptions were issued in year 0 among those participants that later developed dementia. In models where the predictor ACB violated the assumption of linearity between the predictor and the log-hazard (**Suppl. Figure 2**), the covariates were transformed, and the type of transformation is indicated in the results. When a distinct model was run for each predictor, the Bonferroni correction was used. When all predictors were included in a single model, no adjustment for multiple comparisons was done. Results are reported as standardised effect sizes. All analyses were performed in R version 4.1.0 and Python 3.7.10. The code is available at https://github.com/JuM24/UKB-ACB-dementia.

## Results

### Characteristics of the sample

The final sample consisted of 171,775 participants, with diagnoses of dementia dating between July 2002 and June 2020. Among the participants, 2,124 (1.2%) were diagnosed with dementia. The median age of participants at year 0 was 55 years (Q1=49, Q3=59, IQR=10) and the median age of diagnosis with dementia was 72.6 years (IQR=7.2) The characteristics of variables for year 0 are presented in **Table 1** and **Suppl. Table 4**. Depending on the scale used, anticholinergic drugs constituted between 2.5% and 21.8% of all prescriptions between the years 2000 and 2015, with 0.24-2.12 anticholinergic prescriptions per person in year 0 (**Suppl. Table 5**). The characteristics of anticholinergic prescribing in UK Biobank have been described in greater detail elsewhere (Mur et al., DOI:10.1101/2020.10.16.20213884).

**Table 1:**
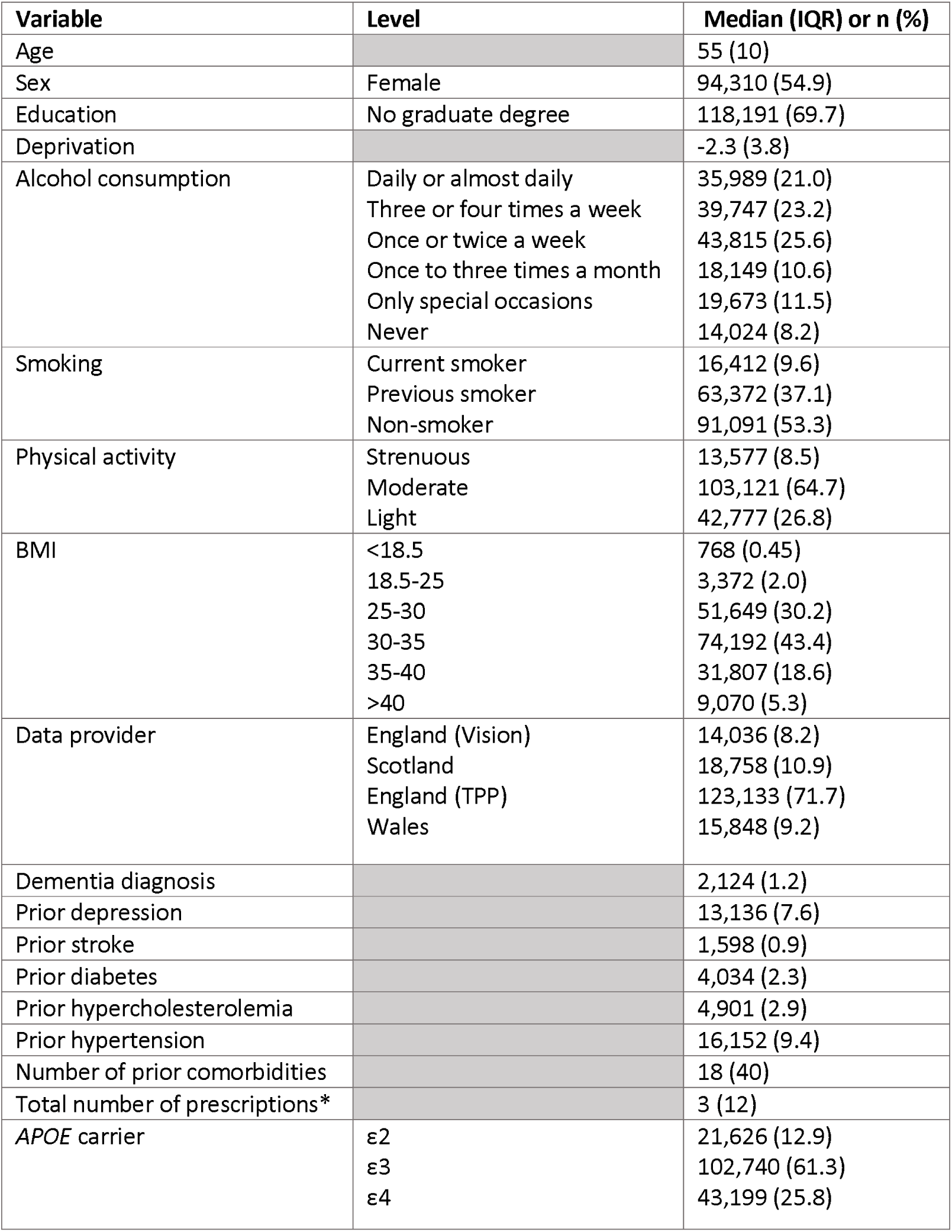
Descriptive statistics of variables used in the models.*The total number of prescriptions was used along the number of anticholinergic drugs to calculate the scale-specific non-anticholinergic drug count.

| Variable | Level | Median (IQR) or n (%) |
| --- | --- | --- |
| Age |  | 55 (10) |
| Sex | Female | 94,310 (54.9) |
| Education | No graduate degree | 118,191 (69.7) |
| Deprivation |  | -2.3 (3.8) |
| Alcohol consumption | Daily or almost daily | 35,989 (21.0) |
|  | Three or four times a week | 39,747 (23.2) |
|  | Once or twice a week | 43,815 (25.6) |
|  | Once to three times a month | 18,149 (10.6) |
|  | Only special occasions | 19,673 (11.5) |
|  | Never | 14,024 (8.2) |
| Smoking | Current smoker | 16,412 (9.6) |
|  | Previous smoker | 63,372 (37.1) |
|  | Non-smoker | 91,091 (53.3) |
| Physical activity | Strenuous | 13,577 (8.5) |
|  | Moderate | 103,121 (64.7) |
|  | Light | 42,777 (26.8) |
| BMI | <18.5 | 768 (0.45) |
|  | 18.5-25 | 3,372 (2.0) |
|  | 25-30 | 51,649 (30.2) |
|  | 30-35 | 74,192 (43.4) |
|  | 35-40 | 31,807 (18.6) |
|  | >40 | 9,070 (5.3) |
| Data provider | England (Vision) | 14,036 (8.2) |
|  | Scotland | 18,758 (10.9) |
|  | England (TPP) | 123,133 (71.7) |
|  | Wales | 15,848 (9.2) |
| Dementia diagnosis |  | 2,124 (1.2) |
| Prior depression |  | 13,136 (7.6) |
| Prior stroke |  | 1,598 (0.9) |
| Prior diabetes |  | 4,034 (2.3) |
| Prior hypercholesterolemia |  | 4,901 (2.9) |
| Prior hypertension |  | 16,152 (9.4) |
| Number of prior comorbidities |  | 18 (40) |
| Total number of prescriptions* |  | 3 (12) |
| APOE carrier | ε2 | 21,626 (12.9) |
|  | ε3 | 102,740 (61.3) |
|  | ε4 | 43,199 (25.8) |

### Anticholinergic scales comparison

Most anticholinergic scales showed positive associations with dementia and with greater effect size estimates than for general polypharmacy (**Figure 1**). HRs for standardised ACB ranged from 1.027 to 1.125 (count-based: median=1.087, IQR=0.044; value-based: median=1.087, IQR=0.019; dosage-based: median=1.078, IQR=0.009; **Suppl. Table 6**). The overlap in CIs was substantial both between scales and within scales; similar results were observed for models with log-and rank-inverse normally transformed predictors (**Suppl. Figure 3**). The value-based scale by Duran et al. (2013)^19^ exhibited the strongest association with dementia (**Suppl. Table 7**) and was used in all subsequent analyses. When death was modelled as a competing outcome, one standard deviation increase in ACB was associated with a 12.0% (95% CI: 7.1%-17.2%) increase in the incidence of dementia, and a 6.0% (95% CI: 3.5%-8.5%) increase in the incidence of all-cause mortality.

**Figure 1:**
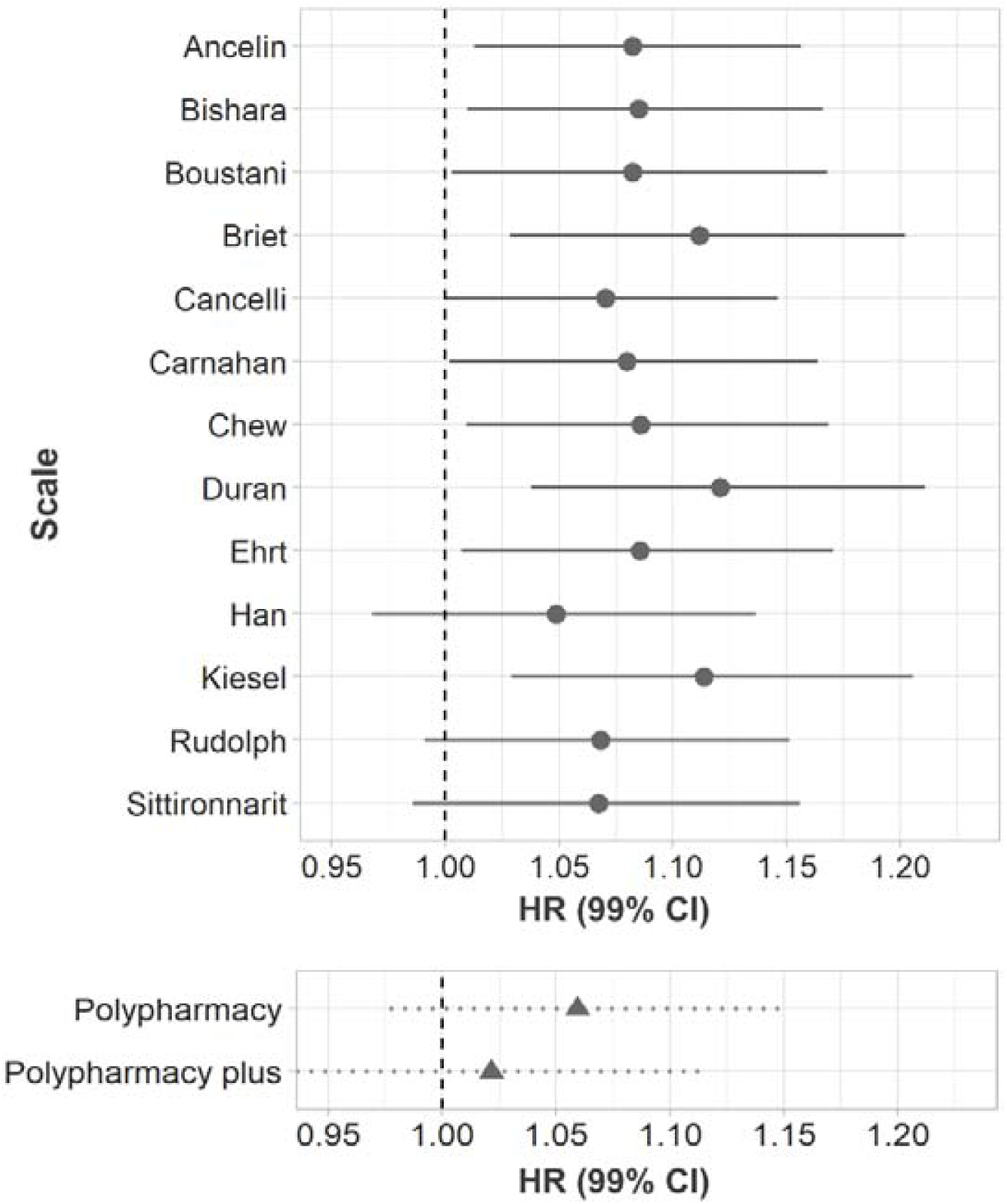
HRs for the association between ACB (top panel) or drug count (bottom panel) and dementia. The names on the y-axis of the top panel refer to the first names of the authors of the original anticholinergic scales; “polypharmacy plus” was additionally controlled for the total number of anticholinergic drugs.

### Time-to-event latency

We compared the risk of dementia occurring within 12 years, between 12-14 years, between 14-16 years, 16-18 years, or more than 18 years (effectively 18-20.5) after year 0. ORs did not differ between most of the different latencies, nor was a pattern discernible in the relationship between latency and effect size (**Table 2**).

**Table 2:** ORs for the risk of dementia within different time periods since the measurement of anticholinergic burden.

| **Latency** (years since 2000) | **OR** | **95% CI** | **N cases** |
| --- | --- | --- | --- |
| 0-12 | 1.20 | 1.05-1.34 | 250 |
| 12-14 | 1.06 | 0.90-1.22 | 257 |
| 14-16 | 1.11 | 1.00-1.22 | 446 |
| 16-18 | 1.21 | 1.10-1.33 | 375 |
| 18-20.5 | 1.07 | 0.98-1.15 | 813 |

### Change in ACB and dementia

The estimate for the association between the individual longitudinal evolution of ACB and dementia was positive (HR=1.094; 95% CI: 1.068-1.119). When the rate of dementia was modelled as a function of the individual longitudinal evolution of ACB in a competing risk model (competing risks: dementia, death), the effect was also positive (death: HR=1.066, 95% CI=1.042-1.089; dementia: HR=1.056, 95% CI=1.008-1.11).

### Drug classes and categories of ACB

Several drug classes exhibited a positive association between ACB and dementia, including drugs for treating the nervous-, gastrointestinal-, and cardiovascular systems (**Figure 2, Suppl. Tables 8,9**). The effect was strongest for antiepileptic drugs, antidepressants, and diuretics (furosemide). While many drugs exhibited a positive tendency for an association between ACB and dementia, the effect sizes were small and the CIs mostly overlapped with HR=1. When the individual yearly drug counts for each group of anticholinergic potency were used to predict dementia (**Figure 3, Suppl. Table 10**), only the number of drugs with an anticholinergic potency of 1 was predictive of dementia.

**Figure 2:**
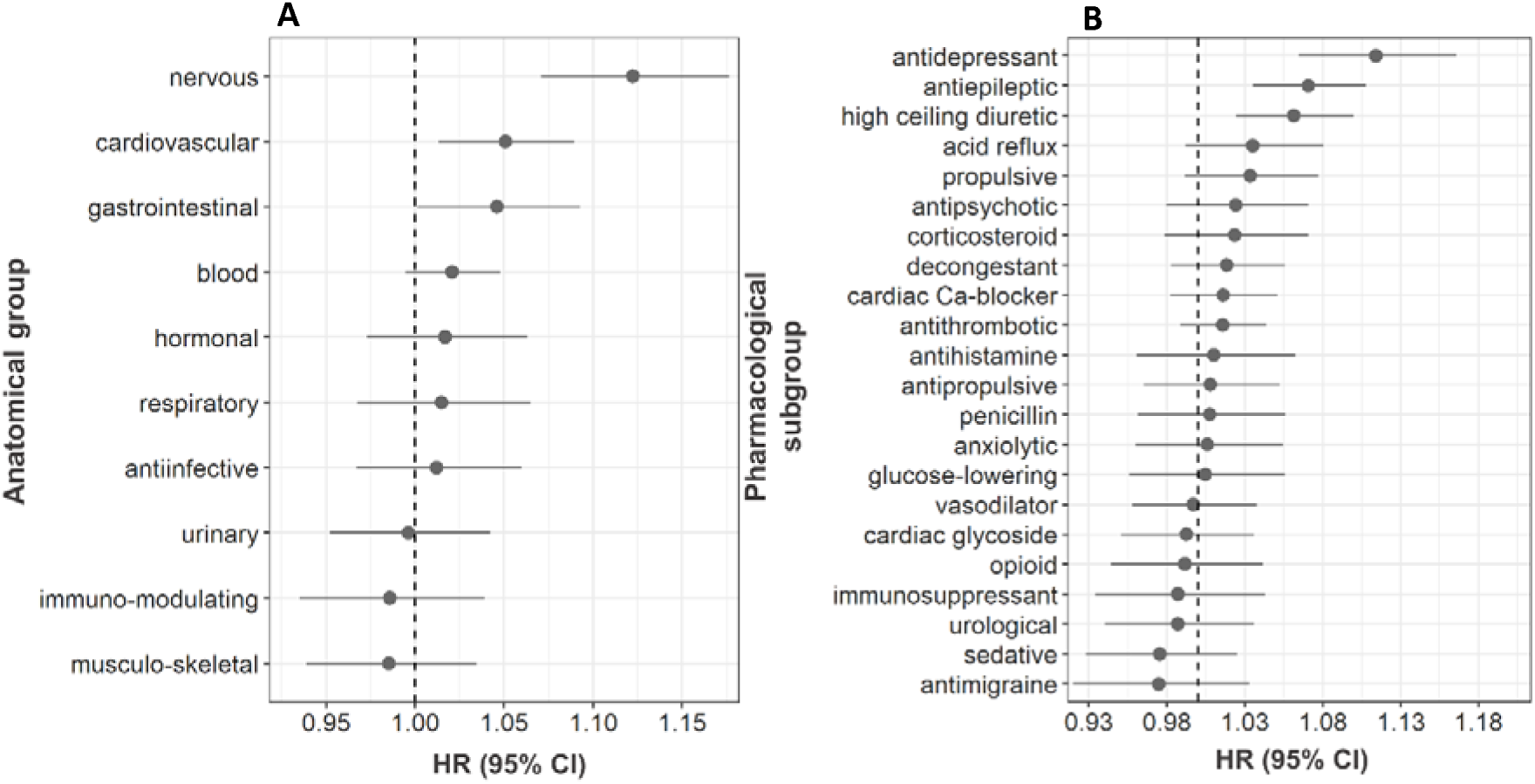
HRs for the association between ACB (rank-based inverse normal transformation) attributable to different classes of drugs and dementia. **A** and **B** reflect the same data, but at different levels of granularity, with **A** representing the topmost level, and **B** the third level from the top according to the WHO classification.

**Figure 3:**
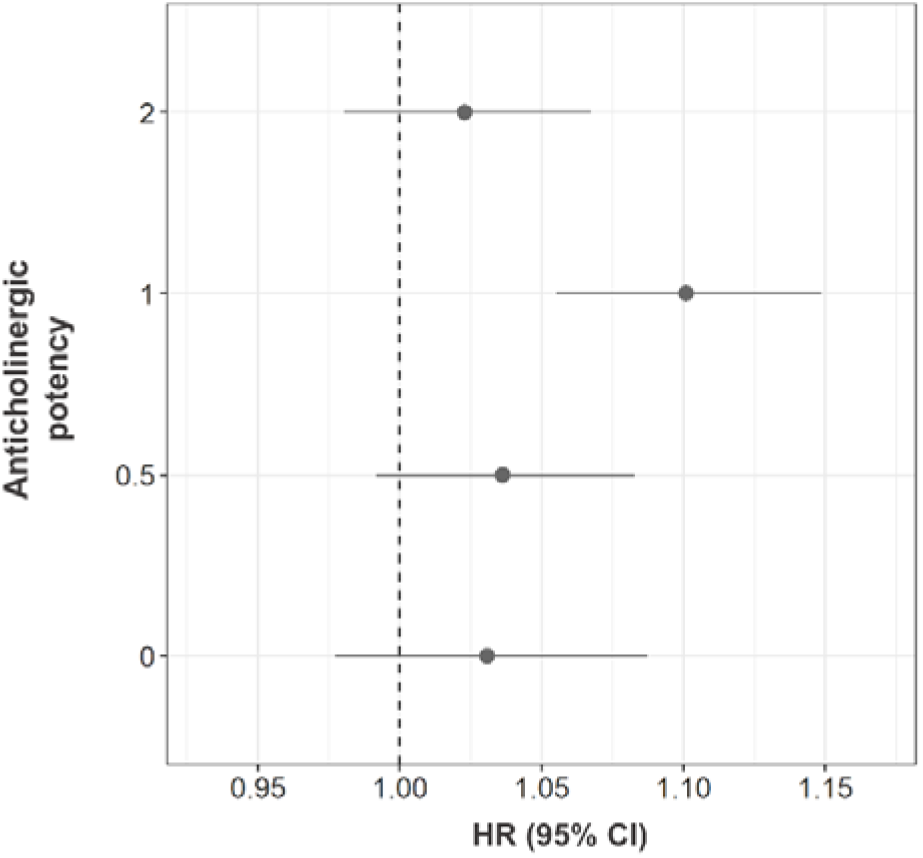
HRs for the association between the numbers of anticholinergic drugs (rank-based inverse normal transformation) of different levels of potency and dementia.

## Discussion

### Interpretation of the findings

In this study, we used electronically prescribed data from 171,775 participants in UK Biobank to study the relationship between ACB and dementia risk. In line with our hypotheses, ACB was associated with dementia across most anticholinergic scales and the best effect estimate for most scales tended to be greater than that for polypharmacy. The data also supported our hypothesis that the trajectory of ACB over time was predictive of dementia, even after accounting for the competing risk of death. The hypotheses regarding class-specific effects were mostly upheld, with ACB due to antidepressants, antiepileptics, antihistamines, and antipsychotics showing positive trends in the association with dementia; however, ACB due to urological drugs did not. We also report associations between additional classes of drugs and risk of dementia, especially high-ceiling diuretics (furosemide). Finally, the strength of the association between ACB and dementia remained unchanged, regardless of the latency between time of measurement and time of diagnosis.

Our results support an association between ACB and dementia across anticholinergic scales, a finding observed before using self-reported medicine use in UK Biobank^30^. This relationship persisted after controlling for several covariates. Across most anticholinergic scales, ACB was a stronger predictor than the total number of prescribed drugs, suggesting that anticholinergic medicines may represent a risk factor distinct from polypharmacy. When applying the anticholinergic scale^19^ that exhibited the strongest association with dementia, ACB also predicted all-cause mortality. Furthermore, not only cumulative ACB measured over one year, but the intra-individual longitudinal trajectory in ACB over the course of 15 years was associated with the risk of dementia. In other words, steeper slopes in the increase of ACB over time were associated with an increased risk of dementia.

However, despite the association between ACB and dementia, several caveats need consideration. First, in contrast to previous findings^5,6,32^ suggesting a dose-response relationship, including dosage into the computation of ACB did not increase model precision or the strength of the association between ACB and dementia. The same was true for the inclusion of anticholinergic scores: simply counting anticholinergic drugs (as opposed to assigning a potency value or weighing by dosage) was equally predictive of dementia. Second, the association between ACB and dementia was limited to ACB attributable to certain classes of drugs. This is consistent with previous findings^5,6^ that reported that ACB attributable to antidepressants, antipsychotics, antihistamines, and antiepileptic drugs was associated with dementia; this consistency was not found for urological drugs, for which the estimate was negative. Third, findings here and elsewhere^6^ indicate that a higher anticholinergic score does not always correspond to a higher risk of dementia.

The consistency in effect sizes for the association between ACB and dementia for different time-to-event latencies has been observed before^5,6^ and suggests that the value of ACB as a potential marker of later cognitive decline does not vary with time. This could indicate the longitudinal consistency in differences in ACB between individuals. While some authors^6^ understand this finding as strengthening the case for causality, it could also – along with the primary finding of an association between ACB and dementia – be explained by confounding by indication: dementia could be caused by the indication for which anticholinergic drugs were prescribed. Indeed, the drugs classes linked to dementia in our study and others^5,6^, are used to treat cardiovascular problems, epilepsy, depression, and schizophrenia, which themselves correlate with neuroanatomical changes or may act as risk factors for dementia^1,33–37^.

### Strengths and weaknesses

The main strengths of our study are the size of the sample, the depth of available data, and the high accuracy of UK Biobank for ascertainment of dementia^38^. Furthermore, our analyses examined ACB from multiple perspectives, including comparing different scales and drug classes. However, we acknowledge several limitations. The participants in UK Biobank are on average healthier and live in less deprived areas than the UK population^39^. Additionally, linked data do not include information on over-the-counter drugs and dietary supplements. Thus, ACB in the UK is likely higher than estimated in our study. Also, due to the low average age of the participants, UK Biobank has relatively few cases of dementia. Next, our analytical approach exhibits weaknesses. First, the quantity/volume of medicines was not used in the calculation of the dosage-based ACB. The validity of our dosage-based scales is thus doubtful. Second, the assumption of linearity between the predictor and the log hazard was sometimes not given and transformations of the data were required to reliably run the models.

### Conclusions and future directions

Inconsistencies in the literature, uncertainty of dose-response-or potency-response relationships, a strong drug-class dependency, and the difficulty to exclude confounding by indication, have led some^40^ to suggest that a different common denominator – other than anticholinergic effect – is responsible for the observed association between anticholinergic drugs and dementia. If correct, the first goal should be the elucidation of the proposed association. Instead of studying the relationship of a general measure of ACB and cognitive decline, researchers could specify and describe the role of distinct classes of anticholinergic medicines – or even individual drugs.

Our study focuses on dementia as an outcome measure, but reports indicate that the associations of anticholinergic use with all-cause mortality^12,41–43^, falls^12,44–46^, and physical function^42^ are also inconsistent. Therefore, we encourage further investigations on these associations on the level of individual drug classes and even individual drugs within the category of anticholinergic drugs.

Considering the role of the cholinergic system in the development of Alzheimer’s disease^3^, a biological underpinning for an effect of anticholinergic drugs on neurodegeneration is intuitive. However, further evidence is needed to determine the brain regions associated with the action of these drugs and the biological pathways likely involved in their proposed effects.

Finally, while previous studies assessed and/or compared anticholinergic scales^12,27,47–50^, questions about their relevance and potential utility remain unanswered. Scales are most often constructed based on expert opinions rooted in past practice and propound established views that might be dated. The contents of anticholinergic scales may certainly reflect a facet of inappropriate prescribing they may help in medical decision-making. However, their heterogeneity and lack of a clear potency-outcome relationship points to an urgent need for reappraisal.

## Data Availability

All the raw data is available to registered researchers via UK Biobank. The code for cleaning the data is available at https://github.com/JuM24/UKB-ACB-dementia.

## Acknowledgements

REM is supported by Alzheimer’s Research UK major project grant ARUK-PG2017B-10. JM is supported by funding from the Wellcome Trust 4-year PhD in Translational Neuroscience—training the next generation of basic neuroscientists to embrace clinical research [108890/Z/15/Z]. JM and TCR are members of the Alzheimer Scotland Dementia Research Centre funded by Alzheimer Scotland. TCR is employed by NHS Lothian and the Scottish Government. SRC is supported by Age UK (Disconnected Mind project), the UK Medical Research Council [MR/R024065/1] and a National Institutes of Health (NIH) research grant R01AG054628.

