## Supplementary material for "Association between anticholinergic burden and dementia in UK Biobank": Suppl. material

**Suppl. Figure 1**: The data cleaning procedure. The grey boxes contain the number of prescriptions (top row in the boxes) and participants (bottom row in the boxes) when a unit of observation was a single prescription; the blue boxes contain the number of prescriptions/participants (both take the same value) when the data was formatted so that the unit of observation was the yearly AB for a participant in year 0. The orange ellipses contain numbers of prescriptions (top row in the ellipses) and participants (bottom row in the ellipses) that were removed at each data-cleaning step. The data cleaning steps include: (1) removal of prescription entries that were blank (i.e., did not list a drug), (2) removal of prescriptions without dates or with invalid dates, (3) the “separation“ of prescriptions with multiple anticholinergic compounds into single entries, (4) removal of prescriptions occurring after the recorded dates of death, (5) removal of prescriptions in years other than year 0, (6) removal of participants diagnoses with dementia prior to year 0 or within one year of year 0, (7) removal of participants diagnosed with Parkinson’s disease, Huntington’s disease, Creutzfeldt-Jacob disease, or multiple sclerosis, (8) removal of participants younger than 60 at the end of sampling or when diagnosed with dementia, (9) removal of participants for whom year 0 was prior to 2015. Please note that in the third step, when prescriptions were “separated” so that prescriptions originally containing several anticholinergic compounds were divided into separate prescriptions (with a single anticholinergic compound each), the number of observations in the dataset effectively *increased*.

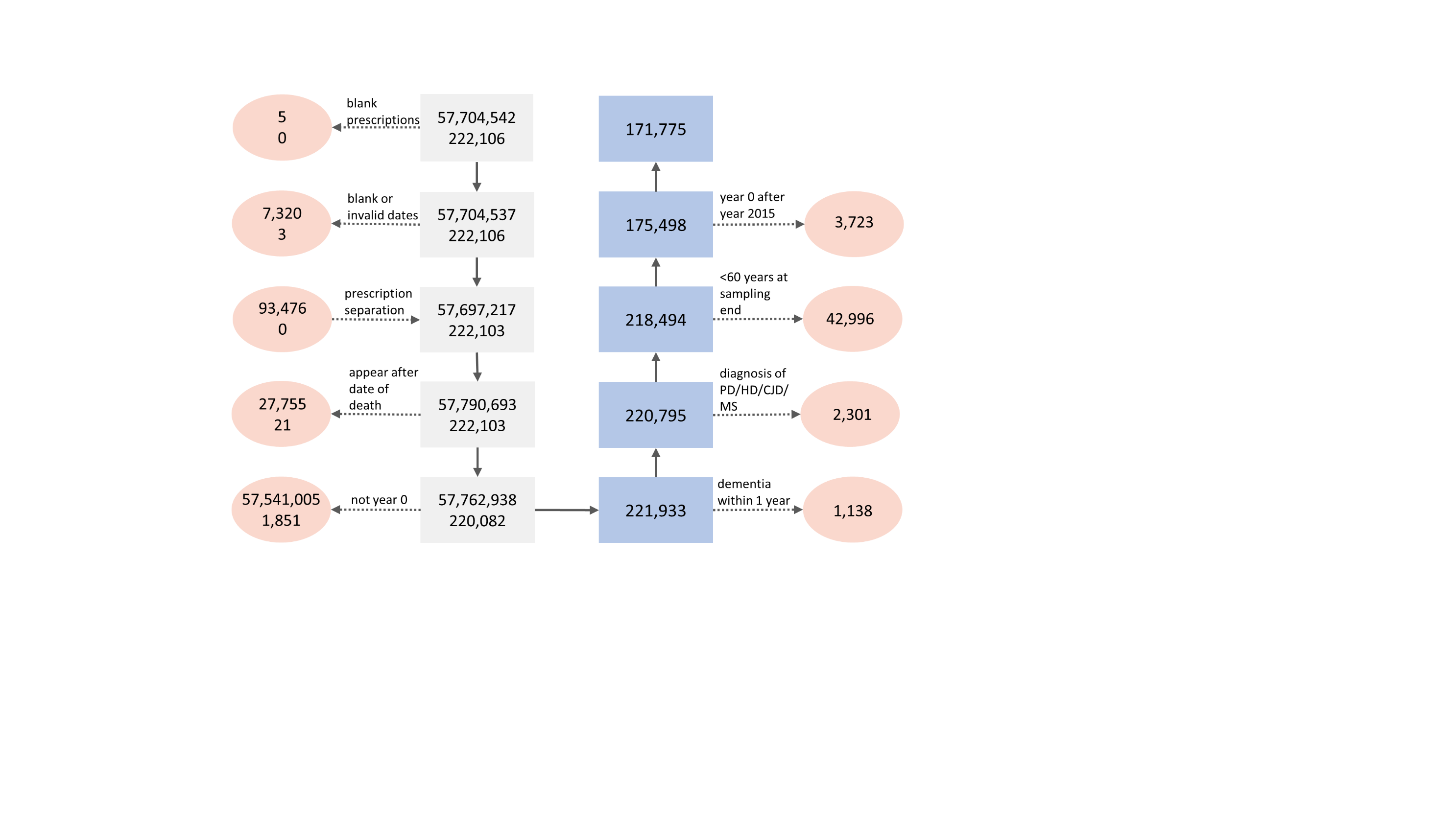

**Suppl. Figure 2**: Martingale residuals plotted against individual continuous covariates. Depicted are only those covariates for which this relationship was judged to not be linear before transforming. For each covariate, four plots are depicted, where the covariate is either untransformed (top left), or square-root-, log- or, rank-based-inverse-normal transformed.

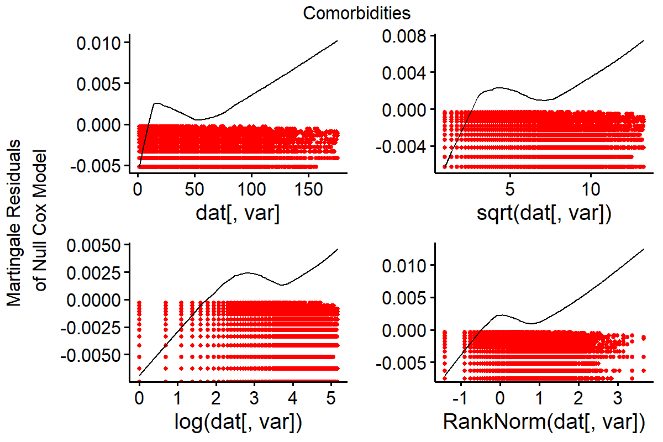

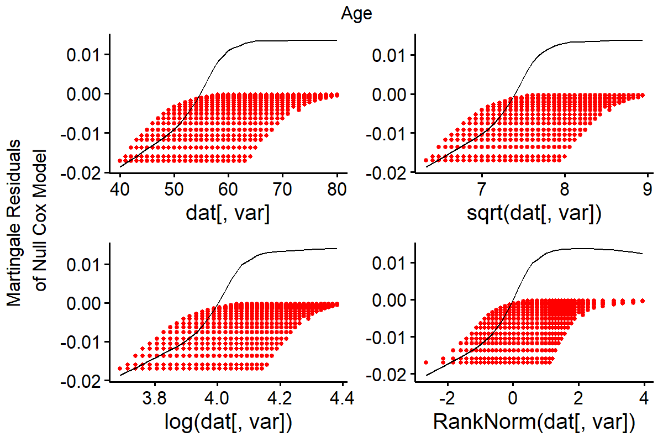

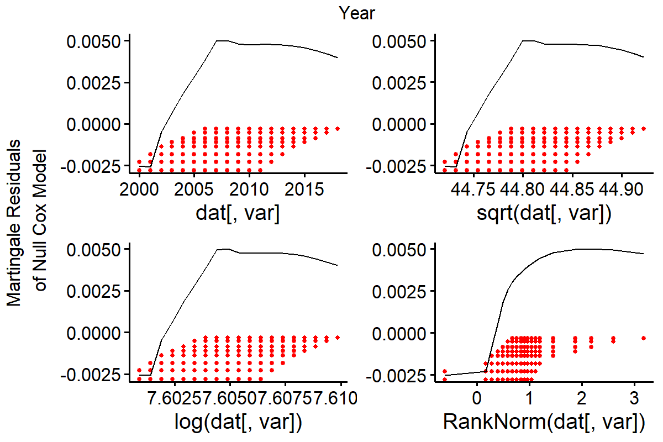

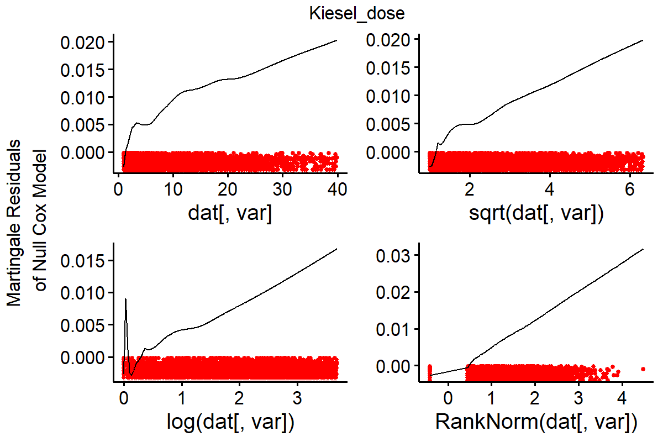

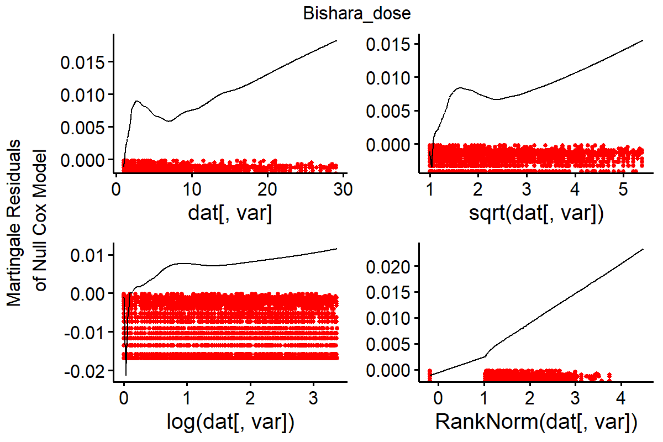

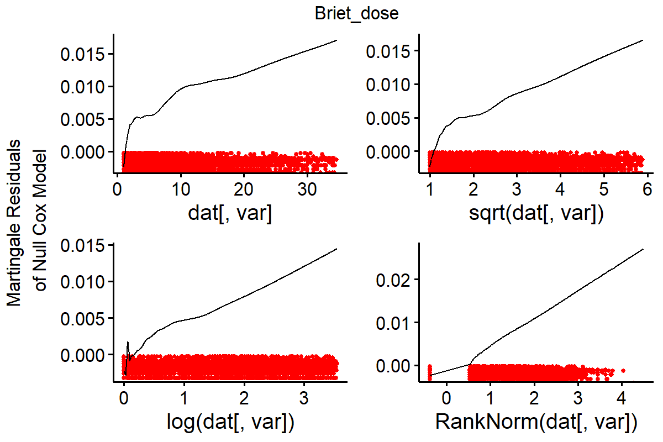

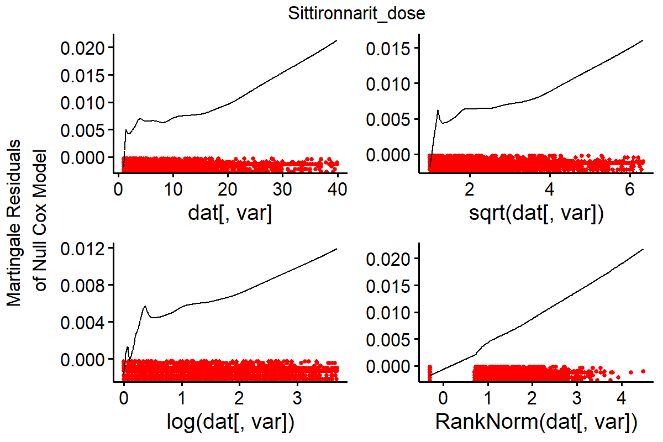

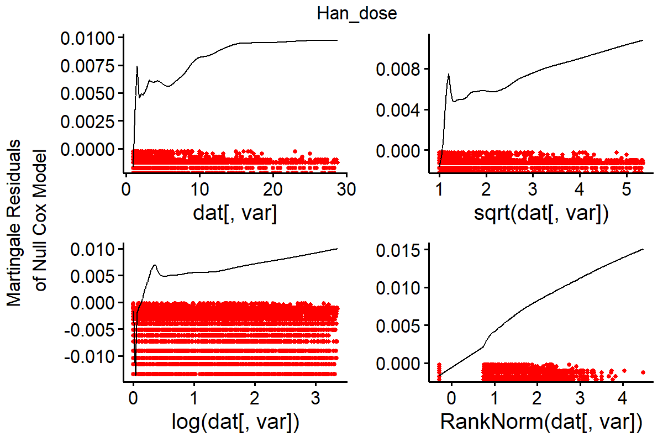

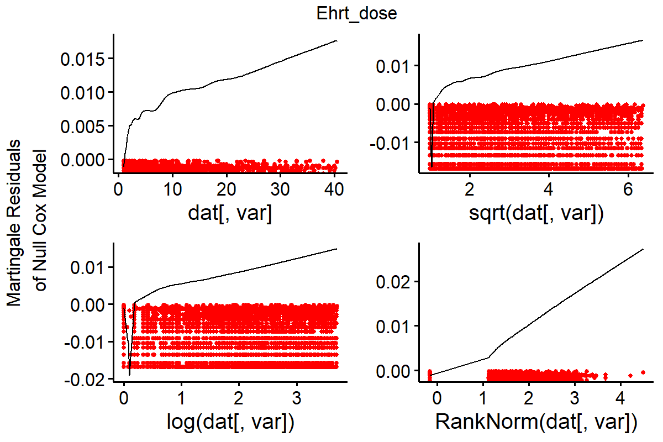

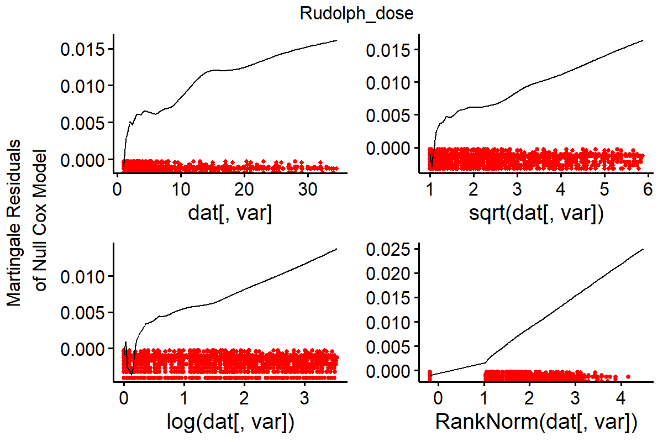

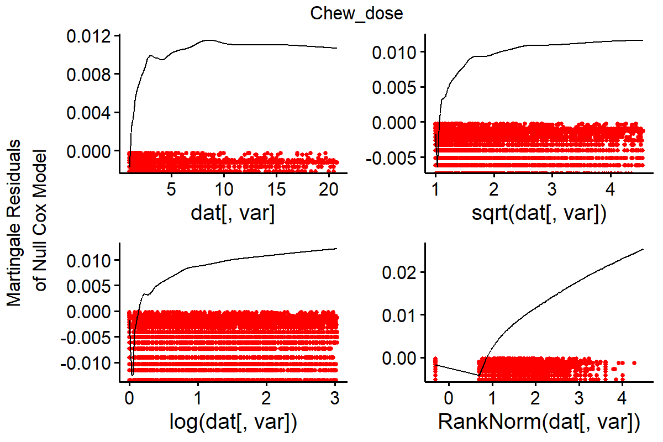

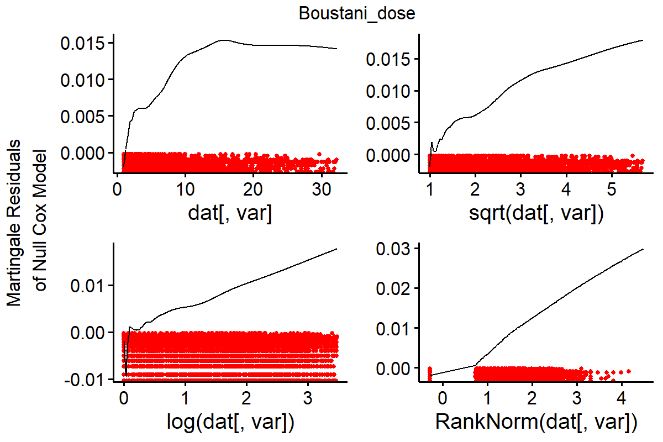

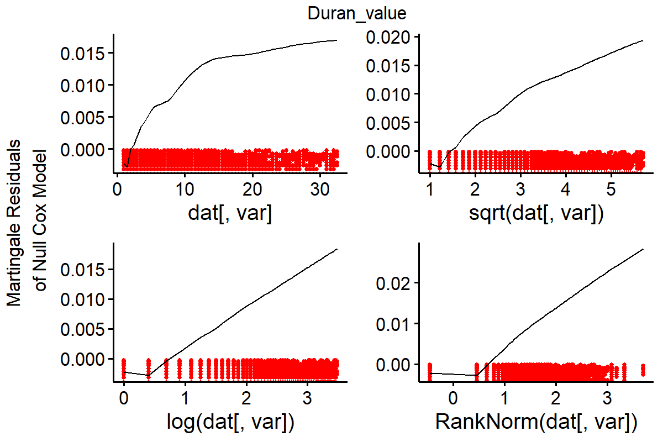

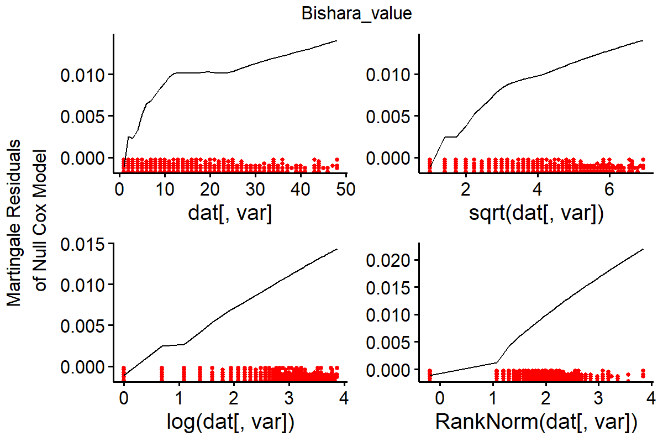

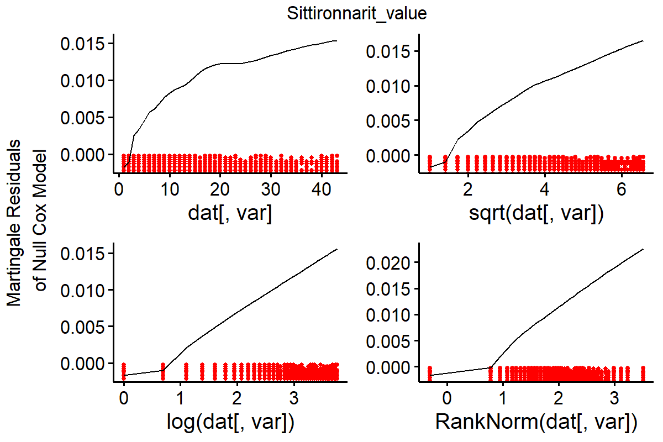

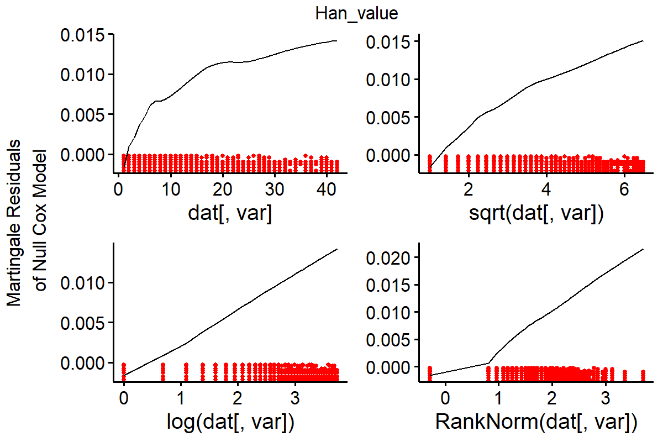

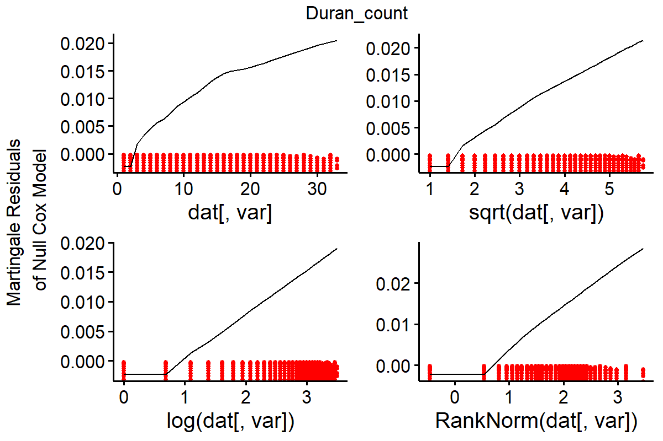

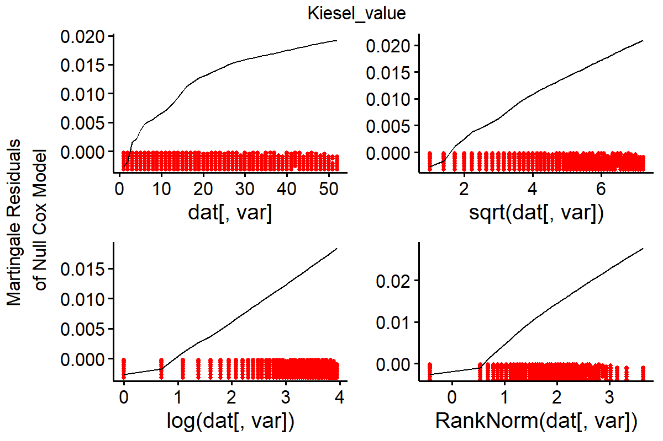

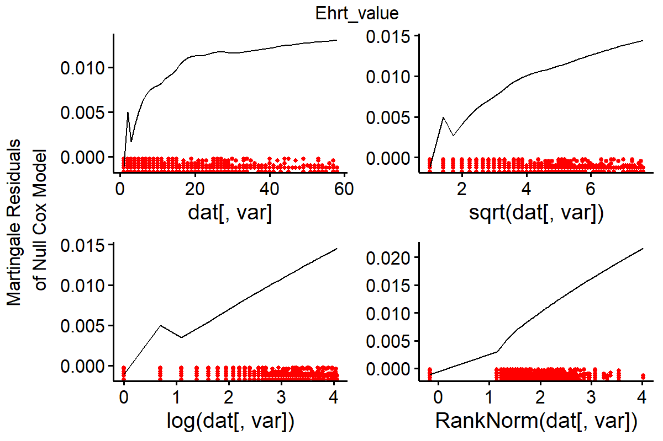

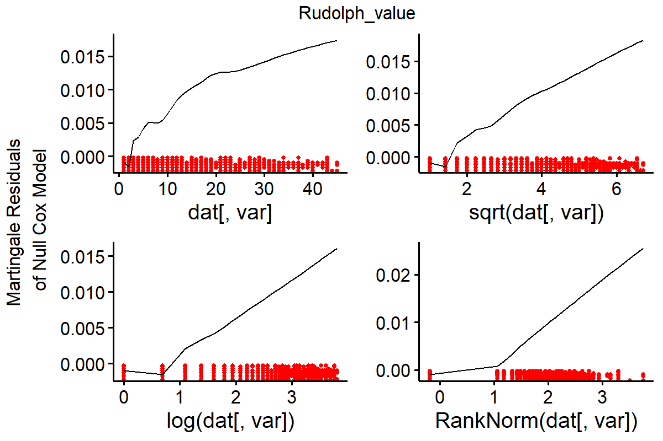

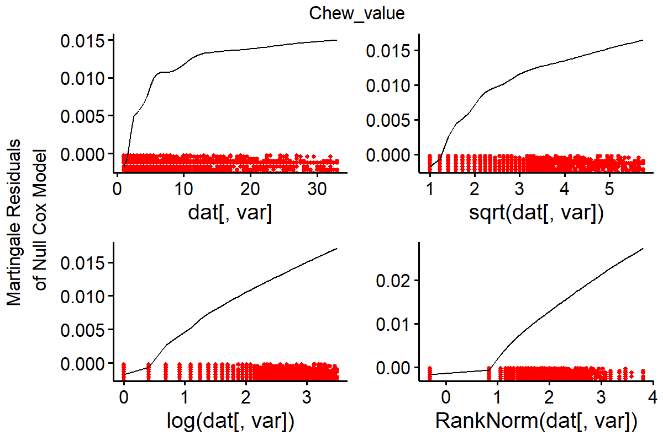

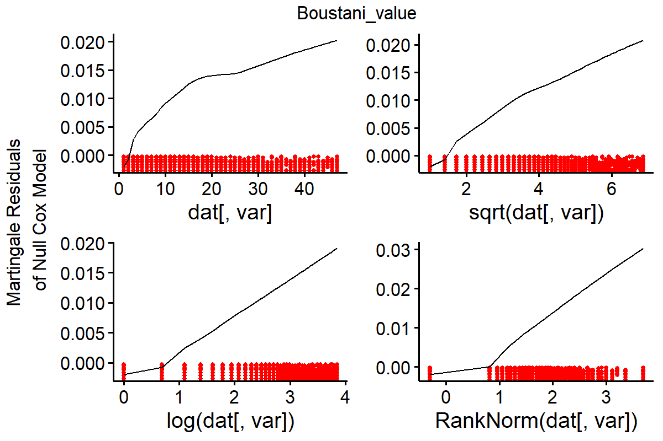

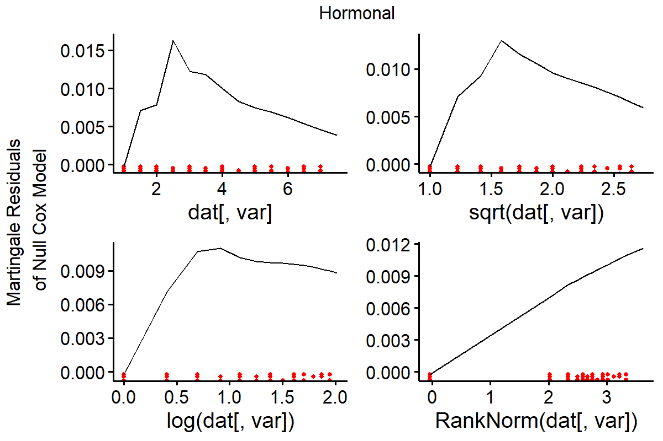

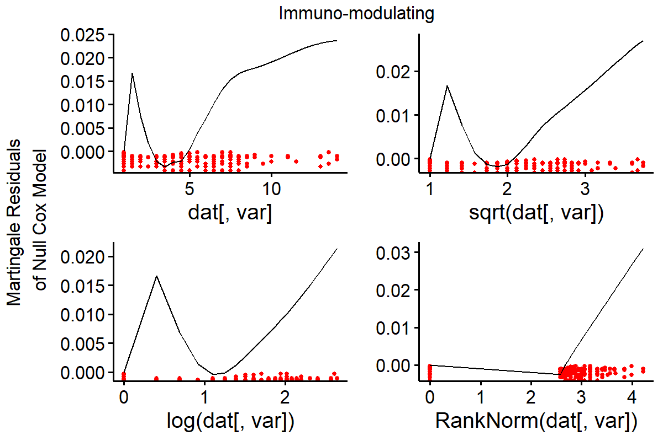

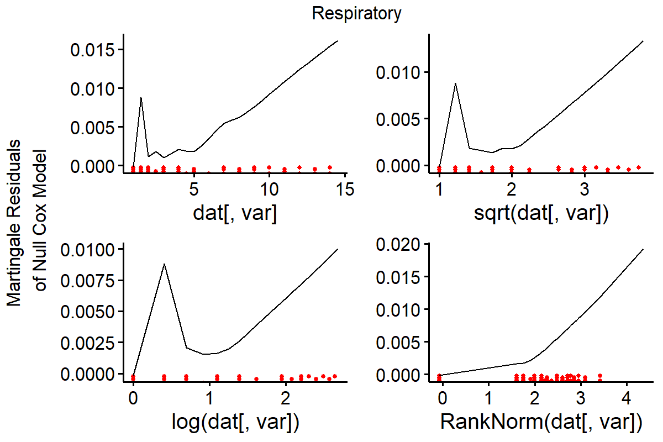

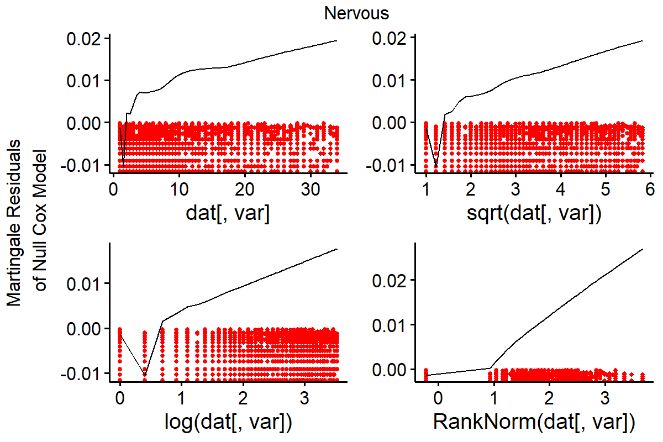

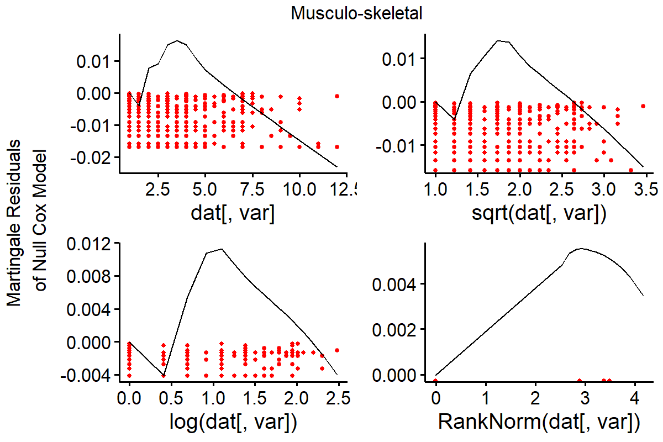

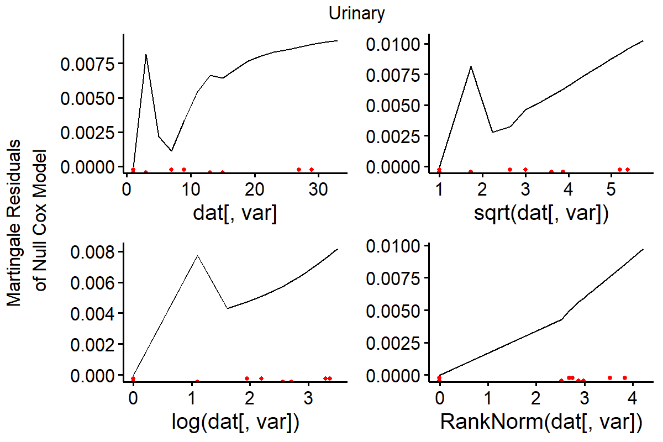

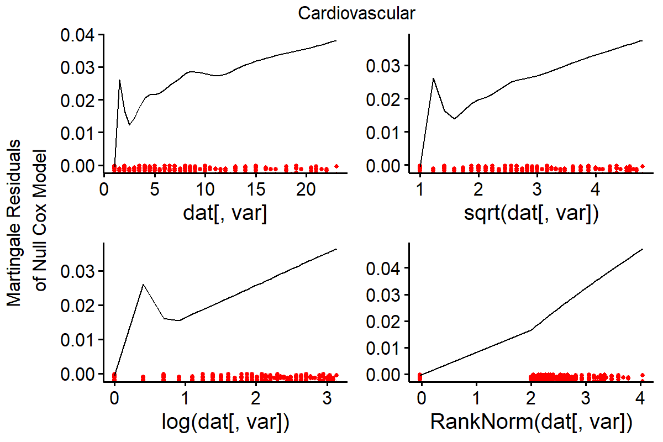

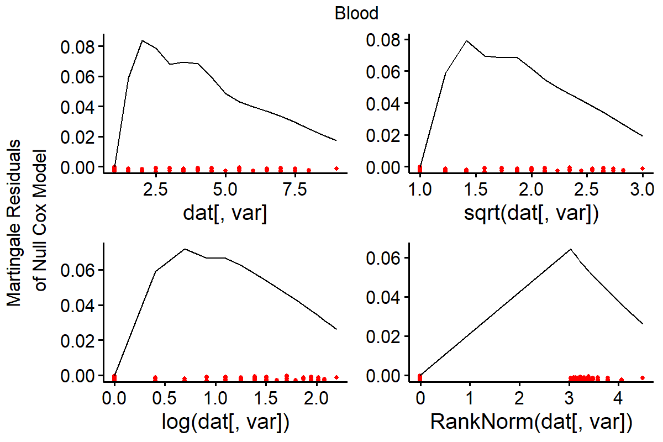

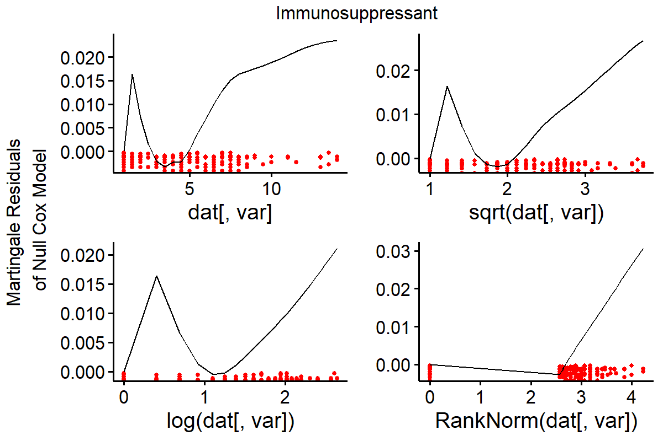

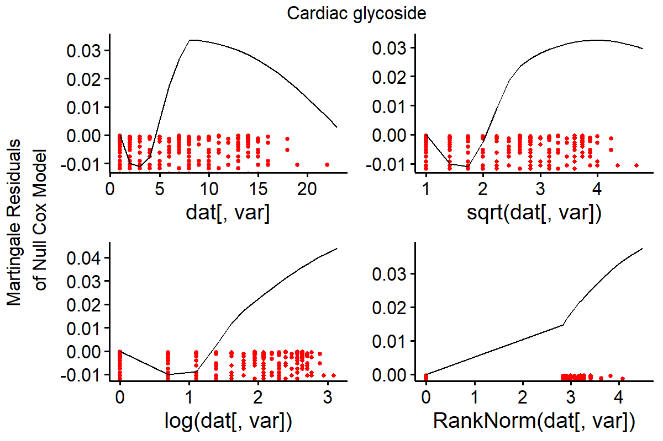

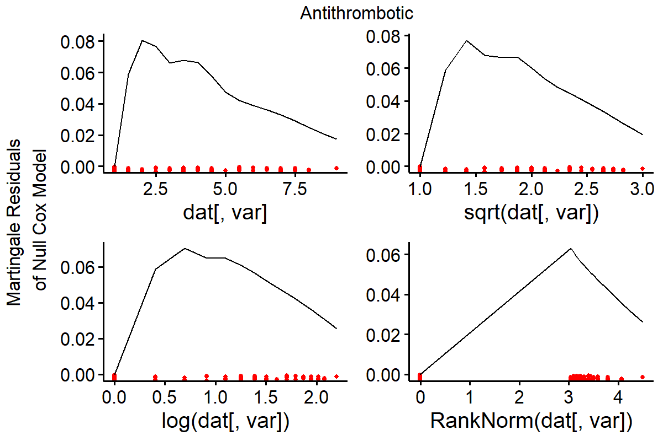

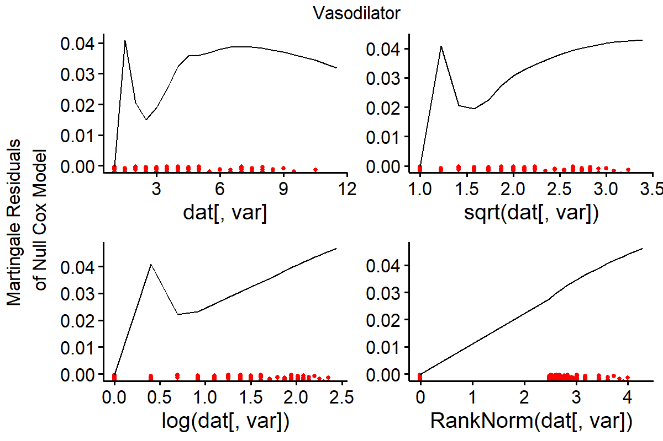

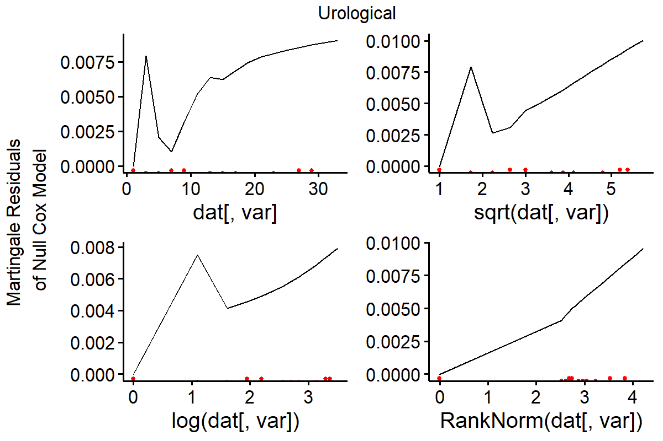

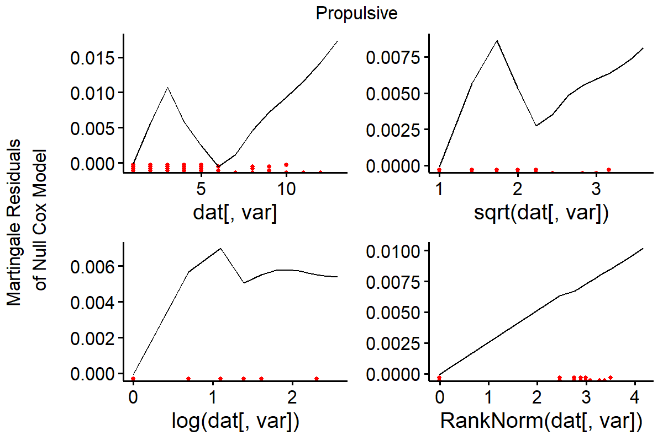

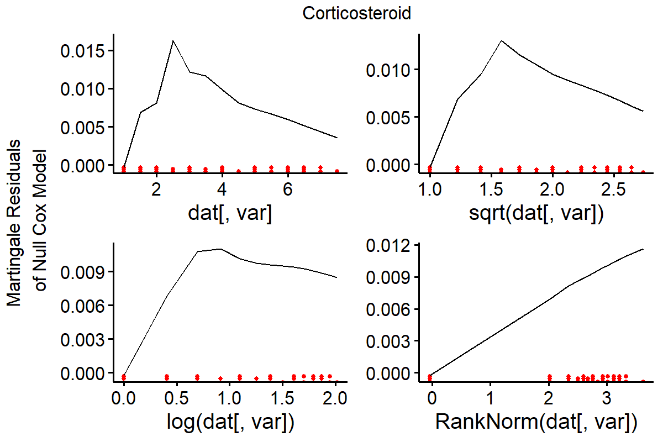

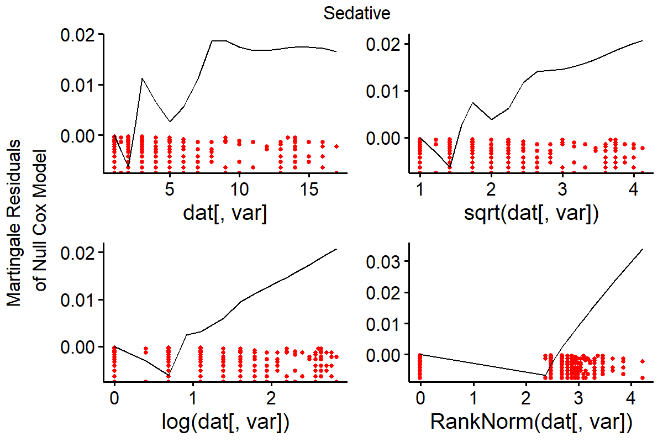

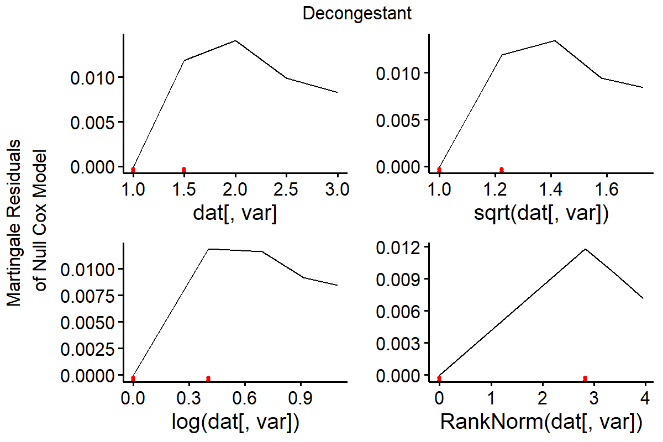

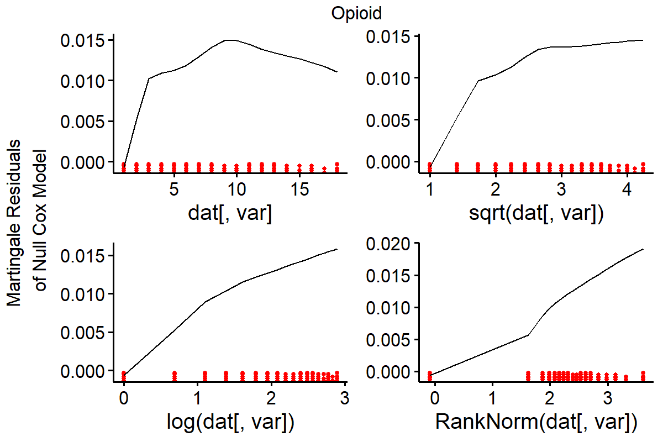

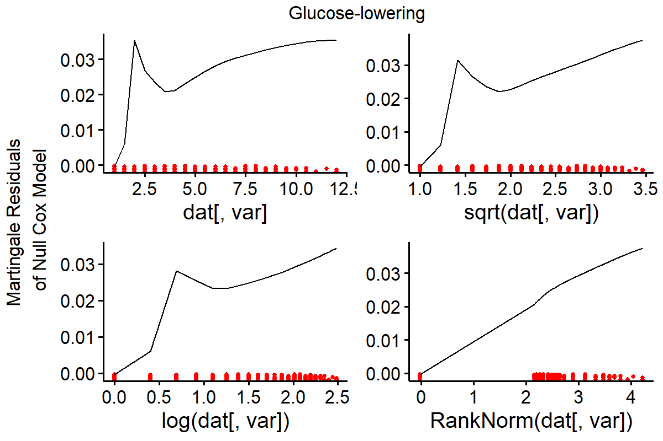

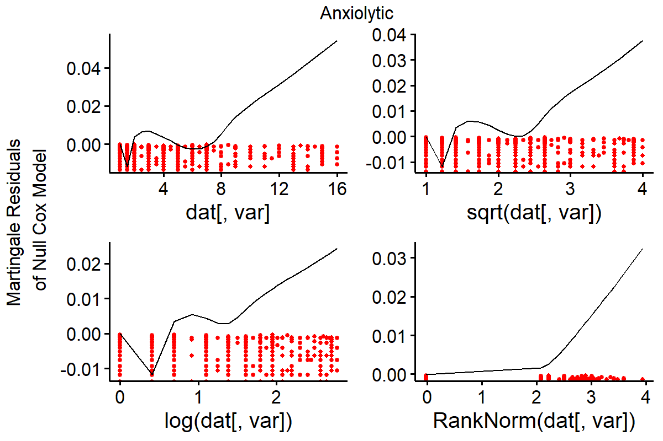

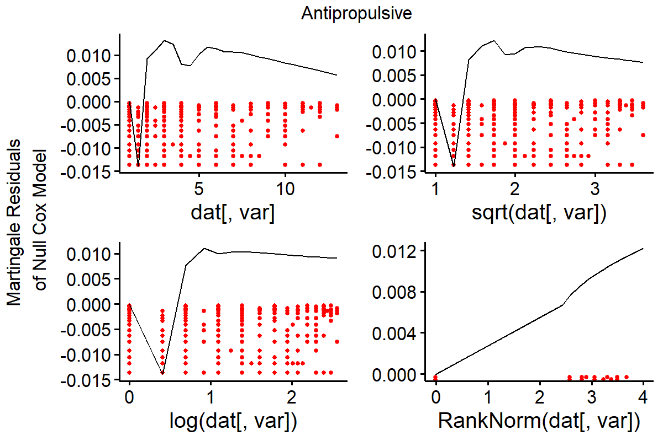

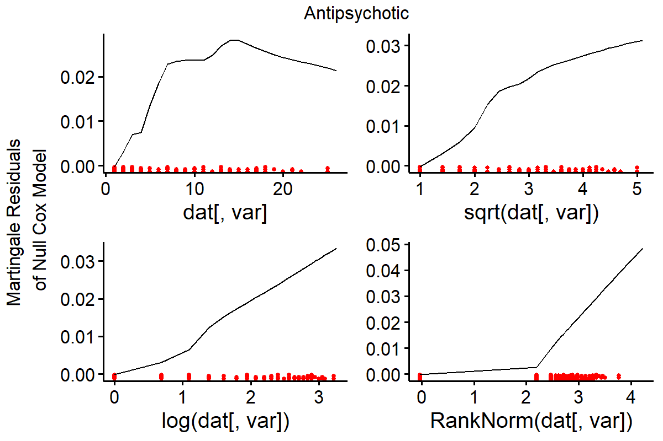

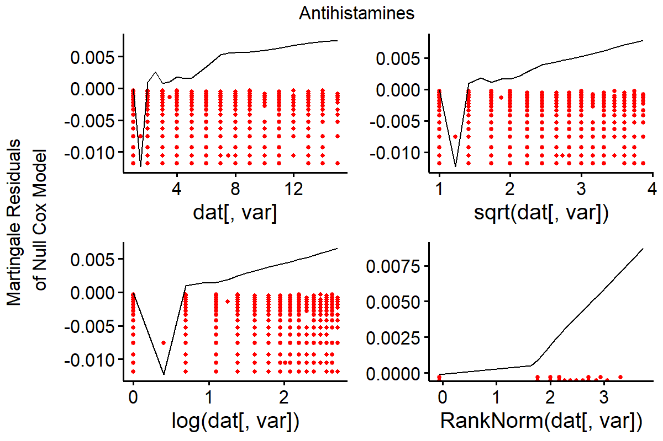

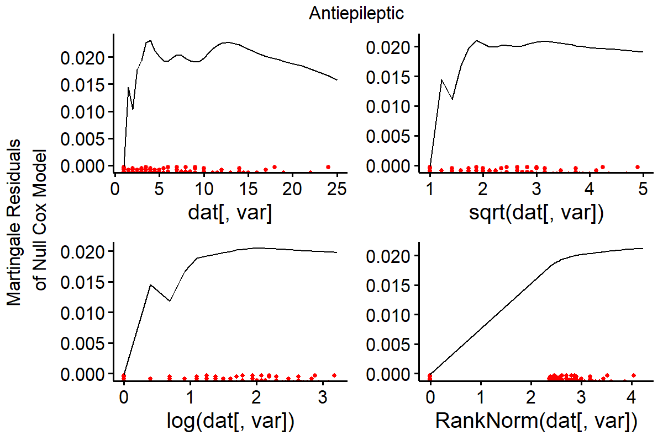

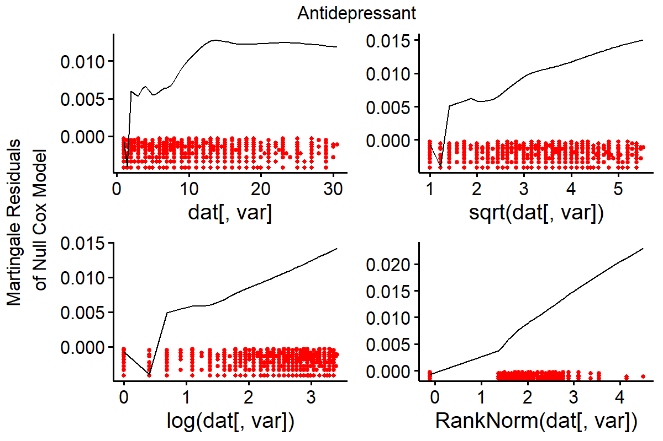

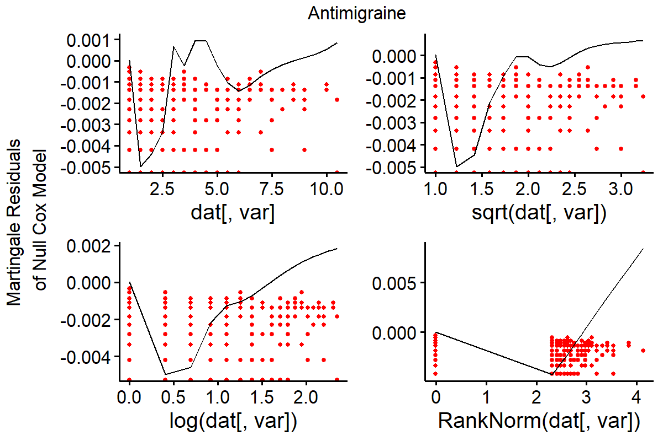

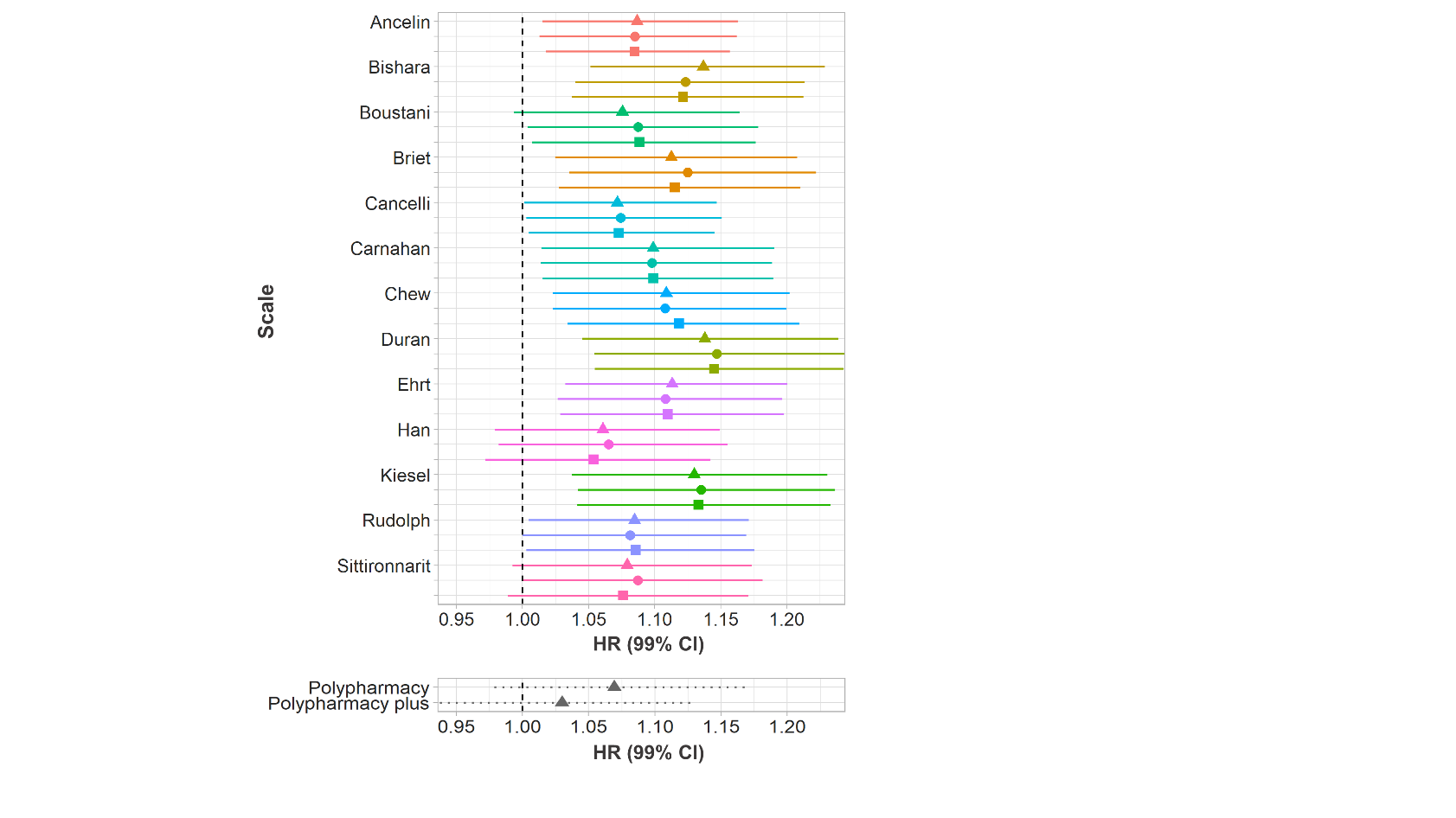

**B**

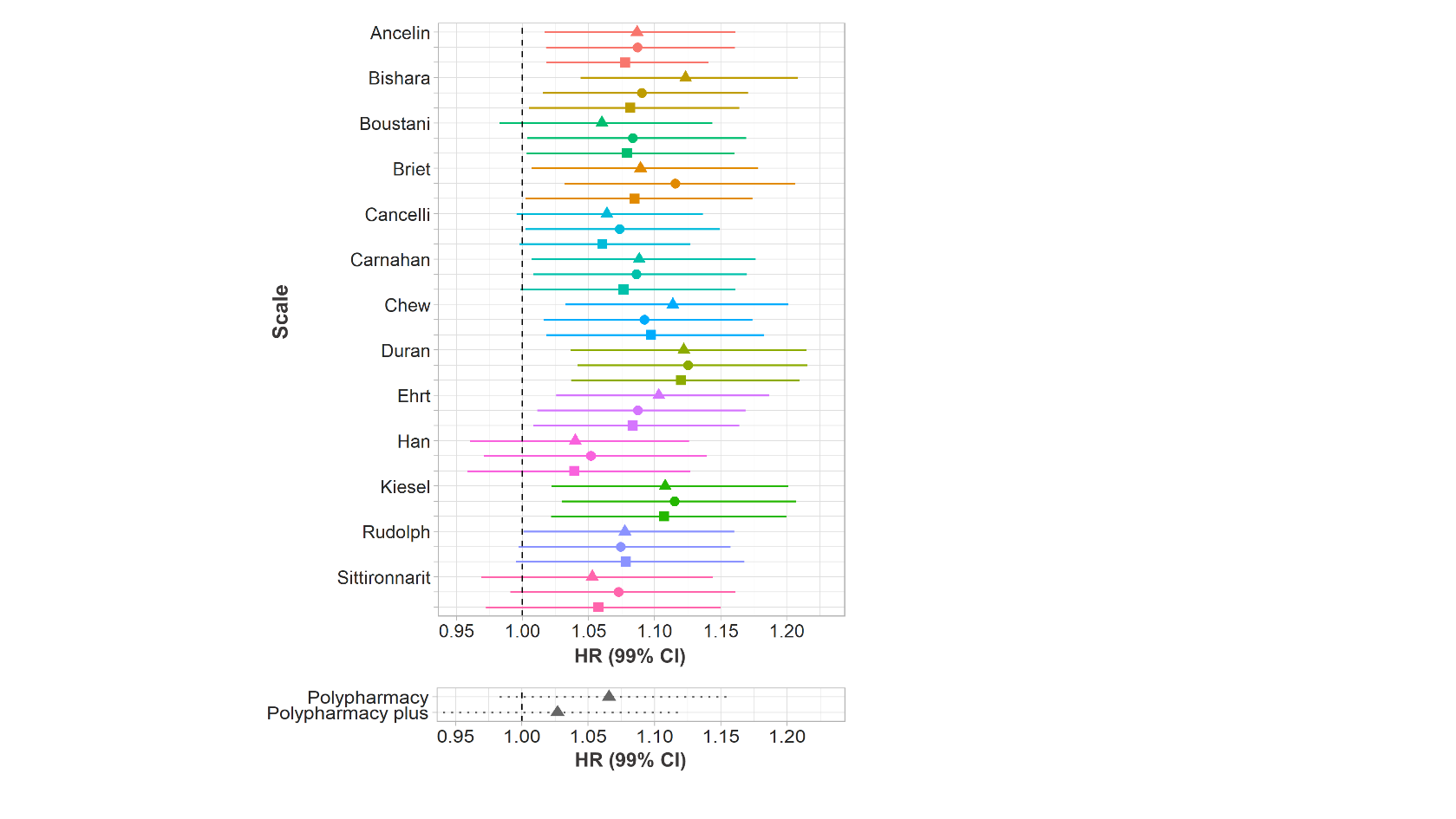

**A**

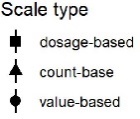

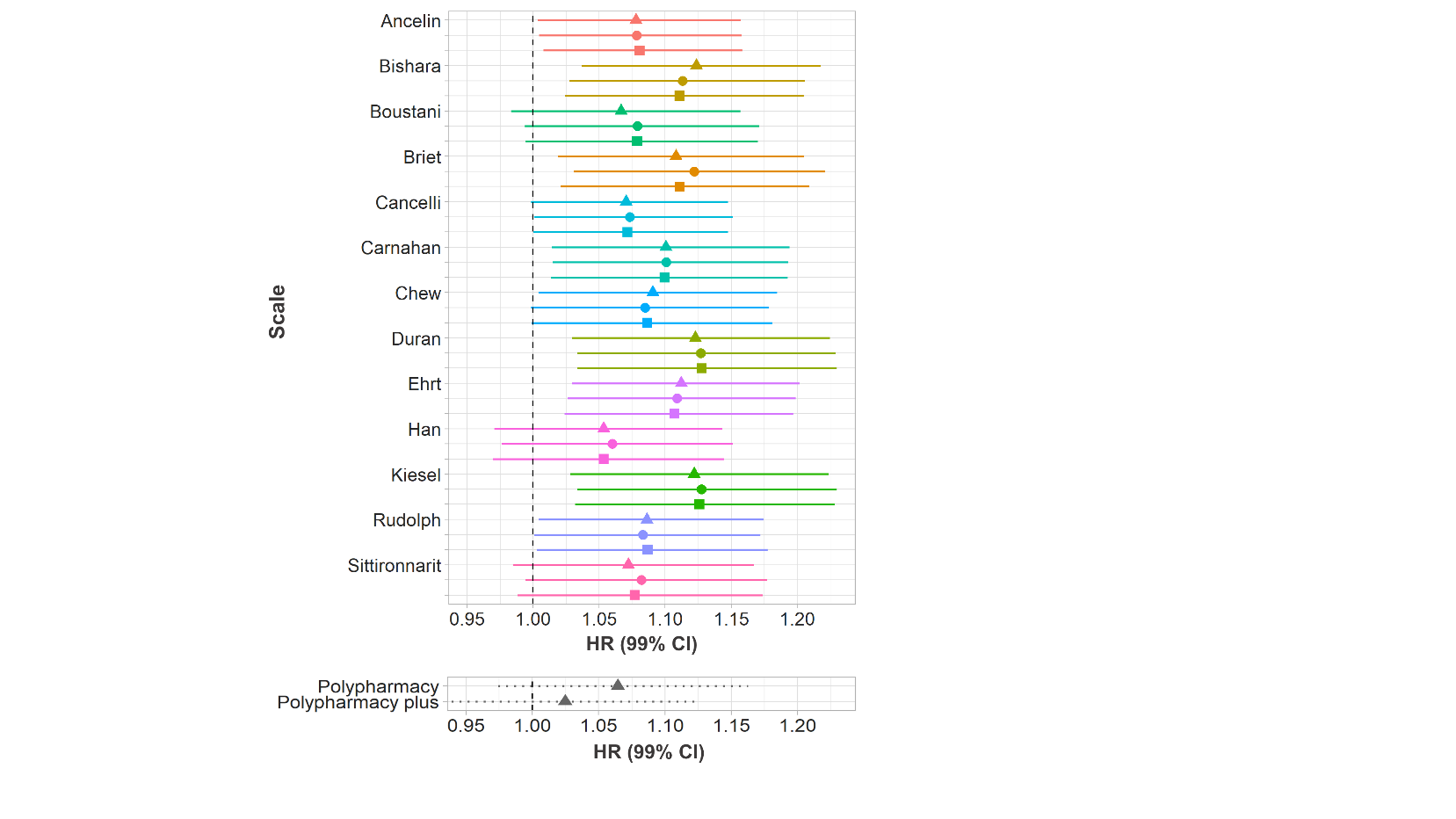

**C**

**Suppl. Figure 3**: HRs for the association between AB and dementia (top panels) and drug count and dementia (bottom panels) when AB is scaled (**A**), log-transformed and scaled (**B**), and transformed using the rank-inverse normal transformation and scaled (**C**). The colour indicates the anticholinergic scales used for the calculation of the AB; the symbols and line types indicate the type of scale computation used.

| **Surname of first author** | **Scale name** | **Year of publication** | **Reason for exclusion** |
| --- | --- | --- | --- |
| Summers^1^ | Drug Risk Number (DRN) | 1978 | Outdated (based on the date of publication and on new scales developed on its basis). |
| Han^2,3^ | Clinician-rated Anticholinergic Scale (CrAS) | 2001 |  |
| Aizenberg^4^ | Anticholinergic Burden Score (ABS) | 2002 | Publicly unavailable and no response from lead author to two email requests within a year. |
| Minzenberg^5^ | n.a. | 2004 | Based on a reference compound. |
| Ancelin^6^ | Anticholinergic Burden Classification (ABC) scale | 2006 |  |
| Carnahan^7^ | Anticholinergic Drug Scale (ADS) | 2006 |  |
| Hilmer^8^ | Drug Burden Index (DBI) | 2007 | Required information on drug dosage. |
| Chew^9^ | Anticholinergic Activity Scale (AAS) | 2008 |  |
| Cancelli^10^ | n.a. | 2008 |  |
| Rudolph^11^ | Anticholinergic Risk Scale (ARS) | 2008 |  |
| Ehrt^12^ | Revised Anticholinergic Activity Scale (AAS-r) | 2010 |  |
| Sittironnarit^13^ | Anticholinergic Loading Scale (ALS) | 2011 |  |
| Boustani^14^ | Anticholinergic Cognitive Burden (ACB) | 2008 |  |
| Whalley^15^ | n.a. | 2012 | Unavailable in full. |
| Durán^16^ | n.a. | 2013 |  |
| Dauphinot^17^ | Drug Burden Index, International Version (DBI-WHO) | 2014 | Required information on drug dosage. |
| Klamer^18^ | MARANTE | 2017 | Required information on drug dosage. |
| Bishara^19^ | Anticholinergic effect on cognition (AEC) scale | 2017 |  |
| Briet^20^ | Anticholinergic impregnation scale | 2017 |  |
| Kiesel^21^ | n.a. | 2018 |  |

**Suppl.** **Table 2**: Anticholinergics scales identified in the present study. We considered anticholinergic scales that were available as complete lists of drugs, scored each drug for its anticholinergic potency, and did not utilize dosage. Grey shading indicates that the scale was not considered for further analysis. For two scales^7,14^, updated versions were used (Aging Brain Care, 2012; Carnahan, 2014, personal communication on 21.10.2019). One scale^11^ was modified to include newer drugs from the UK market as has been done before^22^. Some drugs from one scale^16^ were categorised as “drugs with improbable or no anticholinergic action”. For our analyses, the drugs in that category were scored with 0.5. Modified from Mur et al. (**IN PRESS**).

|  |  | **Dementia diagnosis** | **No dementia diagnosis** |
| --- | --- | --- | --- |
| **Variable** | **Level** | **Median (IQR) or n (%)** | |
| Age |  | 59 (5) | 54 (10) |
| Sex | Female | 1,139 (53.6) | 76,326 (45.0) |
| Education | No graduate degree | 1,672 (80.5) | 116,519 (69.6) |
| Deprivation |  | -1.96 (4.7) | -2.30 (3.8) |
| Alcohol consumption | Daily or almost daily Three or four times a week Once or twice a week Once to three times a month Only special occasions Never | 475 (22.5) 382 (18.1)  461 (21.8) 190 (9.0)  317 (15.0) 289 (13.7) | 35,514 (21.0) 39,365 (23.3)  43,354 (25.6) 17,959 (10.6)  19,356 (11.4) 13,735 (8.1) |
| Smoking | Current smoker  Previous smoker  Non-smoker | 234 (11.1) 935 (44.4) 935 (44.4) | 16,178 (9.6) 62,437 (37.0) 90,156 (53.4) |
| Physical activity | Strenuous  Moderate  Light | 80 (4.3) 1105 (59.9) 661 (35.8) | 13,497 (8.6) 102,016 (64.7) 42,116 (26.7) |
| BMI | <18.5 18.5-25 25-30 30-35 35-40 >40 | 13 (0.62) 606 (28.8) 877 (41.7) 431 (20.5) 129 (6.1) 45 (2.1) | 755 (0.45) 51,043 (30.2) 73,315 (43.4) 31,376 (18.6) 8,941 (5.3) 3,327 (2.0) |
| Data provider | England (Vision) Scotland England (TPP) Wales | 150 (7.1) 40 (1.9) 1,852 (87.2) 82 (3.9) | 13,886 (8.2) 18,718 (11.0) 121,281 (71.5) 15,766 (9.3) |
| Dementia diagnosis |  | 2,124 (100) | 0 (0) |
| Prior depression |  | 257 (12.1) | 12,879 (7.6) |
| Prior stroke |  | 66 (3.1) | 1,532 (0.90) |
| Prior diabetes |  | 184 (8.7) | 3,850 (2.3) |
| Prior hypercholesterolemia |  | 145 (6.8) | 4,756 (2.8) |
| Prior hypertension |  | 415 (19.5) | 15,737 (9.3) |
| Number of prior comorbidities |  | 27 (44) | 18 (40) |
| Total number of prescriptions* |  | 7 (19) | 3 (12) |
| *APOE* carrier | ε2  ε3  ε4 | 191 (7.8) 920 (44.6) 981 (47.6) | 21,465 (13.0) 101,820 (61.5) 41,218 (25.5) |

**Suppl. Table 4**: Descriptive statistics of variables used in the models, presented separately for participants diagnosed with dementia and those not diagnosed with dementia. *The total number of prescriptions was used along the number of anticholinergic drugs to calculate the scale-specific non-anticholinergic drug count.

**Suppl. Table 5**: Frequency of anticholinergic prescribing in the sample from 2000 to 2015 and in year 0 according to each anticholinergic scale studied.

| **Scale** | **Number of distinct anticholinergic drugs in the sample** | **Number of anticholinergic drugs  (% of prescriptions)** | **Number of distinct anticholinergic drugs in the sample in year 0** | **Number of anticholinergic drugs in year 0 (% of prescriptions)** | **Anticholinergic prescriptions per person in year 0** |
| --- | --- | --- | --- | --- | --- |
| Ancelin | 21 | 1,086,739 (2.5) | 20 | 42,068 (2.6) | 0.24 |
| Bishara | 58 | 2,876,150 (6.6) | 56 | 126,195 (7.9) | 0.72 |
| Boustani | 90 | 5,272,868 (12.2) | 87 | 225,409 (14.1) | 1.28 |
| Briet | 121 | 7,041,395 (16.3) | 117 | 294,881 (18.4) | 1.68 |
| Cancelli | 14 | 1,700,948 (3.9) | 13 | 63,609 (4.0) | 0.36 |
| Carnahan | 111 | 3,761,500 (8.7) | 105 | 165,838 (10.4) | 0.94 |
| Chew | 36 | 4,739,876 (11.0) | 34 | 177,101 (11.1) | 1.00 |
| Durán | 147 | 8,257,133 (19.1) | 139 | 320,792 (20.0) | 1.83 |
| Ehrt | 24 | 3,079,302 (7.1) | 23 | 119,600 (7.5) | 0.68 |
| Han | 54 | 4,378,190 (10.1) | 52 | 193,108 (12.1) | 1.10 |
| Kiesel | 141 | 9,495,193 (22.0) | 136 | 371,757 (23.2) | 2.12 |
| Rudolph | 61 | 2,201,774 (5.1) | 59 | 102,540 (6.4) | 0.58 |
| Sittironnarit | 47 | 5,129,912 (11.9) | 46 | 207,085 (12.9) | 1.18 |

| **Scale** | **Type** | **Untransformed** | | **Log** | | **Rank-based inverse-normal** | | **n missing** |
| --- | --- | --- | --- | --- | --- | --- | --- | --- |
|  |  | **HR** | **99% CI** | **HR** | **99% CI** | **HR** | **99% CI** |  |
| Ancelin | count | 1.081 | 1.010-1.156 | 1.081 | 1.009-1.159 | 1.081 | 1.009-1.159 | 23,367 |
|  | value | 1.081 | 1.011-1.156 | 1.080 | 1.007-1.158 | 1.080 | 1.007-1.158 | 23,371 |
|  | dosage | 1.073 | 1.012-1.138 | 1.080 | 1.012-1.153 | 1.080 | 1.012-1.153 | 23,934 |
| Bishara | count | 1.119 | 1.040-1.205 | 1.134 | 1.049-1.227 | 1.134 | 1.049-1.227 | 23,472 |
|  | value | 1.086 | 1.011-1.168 | 1.121 | 1.038-1.212 | 1.121 | 1.038-1.212 | 23,553 |
|  | dosage | 1.079 | 1.002-1.162 | 1.120 | 1.035-1.211 | 1.120 | 1.035-1.211 | 24,130 |
| Boustani | count | 1.053 | 0.975-1.138 | 1.071 | 0.989-1.160 | 1.071 | 0.989-1.160 | 23,525 |
|  | value | 1.077 | 0.996-1.163 | 1.083 | 0.999-1.174 | 1.083 | 0.999-1.174 | 23,698 |
|  | dosage | 1.075 | 0.998-1.157 | 1.084 | 1.003-1.173 | 1.084 | 1.003-1.173 | 24,424 |
| Briet | count | 1.088 | 1.005-1.177 | 1.110 | 1.022-1.206 | 1.110 | 1.022-1.206 | 23,606 |
|  | value | 1.115 | 1.031-1.206 | 1.123 | 1.033-1.221 | 1.123 | 1.033-1.221 | 23,784 |
|  | dosage | 1.085 | 1.003-1.175 | 1.114 | 1.026-1.210 | 1.114 | 1.026-1.210 | 24,327 |
| Cancelli | count | 1.057 | 0.987-1.131 | 1.065 | 0.994-1.141 | 1.065 | 0.994-1.141 | 23,387 |
|  | value | 1.066 | 0.994-1.143 | 1.068 | 0.996-1.145 | 1.068 | 0.996-1.145 | 23,399 |
|  | dosage | 1.055 | 0.991-1.123 | 1.067 | 0.998-1.140 | 1.067 | 0.998-1.140 | 23,966 |
| Carnahan | count | 1.084 | 1.002-1.173 | 1.096 | 1.011-1.188 | 1.096 | 1.011-1.188 | 23,464 |
|  | value | 1.082 | 1.004-1.167 | 1.095 | 1.011-1.186 | 1.095 | 1.011-1.186 | 23,571 |
|  | dosage | 1.075 | 0.996-1.160 | 1.097 | 1.013-1.188 | 1.097 | 1.013-1.188 | 24,204 |
| Chew | count | 1.108 | 1.026-1.196 | 1.105 | 1.019-1.198 | 1.105 | 1.019-1.198 | 23,497 |
|  | value | 1.092 | 1.016-1.174 | 1.107 | 1.022-1.199 | 1.107 | 1.022-1.199 | 23,841 |
|  | dosage | 1.093 | 1.013-1.179 | 1.114 | 1.030-1.206 | 1.114 | 1.030-1.206 | 24,447 |
| Durán | count | 1.120 | 1.034-1.213 | 1.136 | 1.043-1.237 | 1.136 | 1.043-1.237 | 23,559 |
|  | value | 1.124 | 1.040-1.215 | 1.145 | 1.053-1.246 | 1.145 | 1.053-1.246 | 23,768 |
|  | dosage | 1.118 | 1.034-1.208 | 1.143 | 1.052-1.241 | 1.143 | 1.052-1.241 | 24,319 |
| Ehrt | count | 1.096 | 1.018-1.181 | 1.108 | 1.027-1.195 | 1.108 | 1.027-1.195 | 23,412 |
|  | value | 1.082 | 1.005-1.164 | 1.104 | 1.022-1.192 | 1.104 | 1.022-1.192 | 23,485 |
|  | dosage | 1.080 | 1.004-1.161 | 1.106 | 1.024-1.194 | 1.106 | 1.024-1.194 | 24,091 |
| Han | count | 1.040 | 0.960-1.127 | 1.061 | 0.979-1.149 | 1.061 | 0.979-1.149 | 23,577 |
|  | value | 1.052 | 0.971-1.140 | 1.065 | 0.982-1.156 | 1.065 | 0.982-1.156 | 23,730 |
|  | dosage | 1.040 | 0.959-1.128 | 1.054 | 0.972-1.143 | 1.054 | 0.972-1.143 | 24,270 |
| Kiesel | count | 1.105 | 1.019-1.198 | 1.128 | 1.035-1.228 | 1.128 | 1.035-1.228 | 23,561 |
|  | value | 1.113 | 1.027-1.205 | 1.133 | 1.040-1.235 | 1.133 | 1.040-1.235 | 23,750 |
|  | dosage | 1.105 | 1.020-1.198 | 1.131 | 1.039-1.231 | 1.131 | 1.039-1.231 | 24,405 |
| Rudolph | count | 1.077 | 1.000-1.160 | 1.084 | 1.004-1.171 | 1.084 | 1.004-1.171 | 23,487 |
|  | value | 1.072 | 0.995-1.156 | 1.081 | 0.999-1.169 | 1.081 | 0.999-1.169 | 23,529 |
|  | dosage | 1.078 | 0.994-1.168 | 1.085 | 1.002-1.175 | 1.085 | 1.002-1.175 | 24,060 |
| Sittironnarit | count | 1.055 | 0.971-1.146 | 1.081 | 0.994-1.175 | 1.081 | 0.994-1.175 | 23,534 |
|  | value | 1.074 | 0.993-1.163 | 1.089 | 1.002-1.183 | 1.089 | 1.002-1.183 | 23,678 |
|  | dosage | 1.059 | 0.974-1.151 | 1.077 | 0.990-1.172 | 1.077 | 0.990-1.172 | 24,293 |
| Polypharmacy | count | 1.062 | 0.979-1.151 | 1.067 | 0.976-1.165 | 1.067 | 0.976-1.165 | 24,103 |
| Polypharmacy plus | count | 1.025 | 0.939-1.118 | 1.029 | 0.937-1.130 | 1.029 | 0.937-1.130 | 24,103 |

**Suppl. Table 6**: HRs for scaled numerical variables in the Cox proportional risks model predicting the risk of dementia. Each row depicts the effect of anticholinergic burden according to a different anticholinergic scale. The different columns depict HRs for different transformations of the data.

| **Variable** | **Level** | **HR** | **95% CI** |
| --- | --- | --- | --- |
| AB |  | 1.124 | 1.040-1.215 |
| Sex | Female | 1.459 | 1.223-1.742 |
| Year 0 |  | 0.913 | 0.771-1.081 |
| Age 0 |  | 4.266 | 3.672-4.957 |
| Education | Graduate degree | 0.825 | 0.672-1.012 |
| Deprivation |  | 1.084 | 0.997-1.178 |
| Alcohol consumption | Three or four times a week Once or twice a week Once to three times a month Only special occasions Never | 0.834 0.896 0.899 1.127 1.343 | 0.651-1.067 0.706-1.138 0.654-1.237 0.846-1.500 0.994-1.814 |
| Smoking | Current smoker  Previous smoker | 1.127 1.264 | 0.945-1.344 0.937-1.704 |
| Physical activity | Strenuous  Moderate | 0.764 0.657 | 0.639-0.912 0.436-0.991 |
| BMI | <18.5 25-30 30-35 35-40 >40 | 0.646 0.537 0.511 0.552 0.499 | 0.240-1.736 0.200-1.441 0.188-1.389 0.193-1.575 0.147-1.028 |
| Data provider | England TPP Scotland Wales | 1.433 0.236 0.528 | 1.017-2.018 0.119-0.470 0.314-0.887 |
| Prior depression |  | 1.182 | 0.895-1.562 |
| Prior stroke |  | 1.545 | 0.867-2.753 |
| Prior diabetes |  | 1.840 | 1.242-2.726 |
| Prior hypercholesterolemia |  | 1.057 | 0.708-1.579 |
| Prior hypertension |  | 1.249 | 0.977-1.597 |
| Number of prior comorbidities |  | 1.052 | 0.967-1.145 |
| Non-anticholinergic drug count |  | 1.031 | 0.941-1.130 |
| *APOE* carrier | ε2  ε4 | 1.206 3.296 | 0.879-1.655 2.408-4.511 |

**Suppl. Table 7**: HRs for scaled numerical variables in the Cox proportional risks model predicting the risk of dementia. Anticholinergic burden was determined using the value-based scale by Durán et al. (2013)^16^. The CIs have been adjusted for multiple comparisons (n=41).

| **Anatomical group** | **HR** | **95% CI** | **n missing** |
| --- | --- | --- | --- |
| nervous | 1.123 | 1.071-1.178 | 19,700 |
| cardiovascular | 1.052 | 1.015-1.090 | 19,700 |
| gastrointestinal | 1.047 | 1.002-1.094 | 19,700 |
| blood | 1.022 | 0.996-1.049 | 19,700 |
| hormonal | 1.018 | 0.974-1.065 | 19,700 |
| respiratory | 1.010 | 0.962-1.061 | 19,700 |
| antiinfective | 1.010 | 0.965-1.058 | 19,700 |
| urinary | 0.991 | 0.946-1.039 | 19,700 |
| immuno-modulating | 0.986 | 0.936-1.040 | 19,700 |
| musculo-skeletal | 0.985 | 0.938-1.033 | 19,700 |

**Suppl. Table 8**: HRs for scaled numerical variables in the Cox proportional risks model predicting the risk of dementia. Each row depicts the effect of anticholinergic burden due to a drug prescribed for a different anatomical group.

**Suppl. Table 9**: HRs for scaled numerical variables in the Cox proportional risks model predicting the risk of dementia. Each row depicts the effect of anticholinergic burden due to a drug prescribed for a different pharmacological group.

| **Pharmacological group** | **HR** | **95% CI** | **n missing** |
| --- | --- | --- | --- |
| antidepressant | 1.114 | 1.065-1.166 | 20,047 |
| antiepileptic | 1.071 | 1.035-1.108 | 20,047 |
| high ceiling diuretic | 1.061 | 1.024-1.100 | 20,047 |
| acid reflux | 1.035 | 0.992-1.080 | 20,047 |
| propulsive | 1.033 | 0.991-1.077 | 20,047 |
| antipsychotic | 1.024 | 0.980-1.071 | 20,047 |
| corticosteroid | 1.024 | 0.979-1.071 | 20,047 |
| decongestant | 1.018 | 0.982-1.056 | 20,047 |
| cardiac Ca-blocker | 1.016 | 0.982-1.051 | 20,047 |
| antithrombotic | 1.016 | 0.989-1.044 | 20,047 |
| antihistamine | 1.010 | 0.961-1.061 | 20,047 |
| antipropulsive | 1.008 | 0.965-1.052 | 20,047 |
| penicillin | 1.008 | 0.961-1.056 | 20,047 |
| anxiolytic | 1.006 | 0.960-1.054 | 20,047 |
| glucose-lowering | 1.005 | 0.956-1.056 | 20,047 |
| vasodilator | 0.997 | 0.958-1.038 | 20,047 |
| cardiac glycoside | 0.992 | 0.951-1.036 | 20,047 |
| opioid | 0.992 | 0.944-1.042 | 20,047 |
| immunosuppressant | 0.987 | 0.934-1.043 | 20,047 |
| urological | 0.987 | 0.940-1.036 | 20,047 |
| sedative | 0.975 | 0.928-1.025 | 20,047 |
| antimigraine | 0.975 | 0.920-1.033 | 20,047 |

**Suppl. Table 10**: HRs for scaled numerical variables in the Cox proportional risks model predicting the risk of dementia. Each row depicts the effect of anticholinergic burden due to a drug prescribed for a different category of anticholinergic potency.

| **Potency score** | **HR** | **95% CI** | **n missing** |
| --- | --- | --- | --- |
| 2 | 1.026 | 0.984-1.070 | 20,693 |
| 1 | 1.097 | 1.051-1.145 | 20,693 |
| 0.5 | 1.034 | 0.990-1.081 | 20,693 |
| 0 | 1.032 | 0.978-1.089 | 20,693 |
